# Neuroimaging Biomarkers in Addiction

**DOI:** 10.1101/2024.09.02.24312084

**Authors:** Hamed Ekhtiari, Arshiya Sangchooli, Owen Carmichael, F. Gerard Moeller, Patricio O’Donnell, Maria Oquendo, Martin P. Paulus, Diego A. Pizzagalli, Tatiana Ramey, Joseph Schacht, Mehran Zare-Bidoky, Anna Rose Childress, Kathleen Brady

## Abstract

As a neurobiological process, addiction involves pathological patterns of engagement with substances and a range of behaviors with a chronic and relapsing course. Neuroimaging technologies assess brain activity, structure, physiology, and metabolism at scales ranging from neurotransmitter receptors to large-scale brain networks, providing unique windows into the core neural processes implicated in substance use disorders. Identified aberrations in the neural substrates of reward and salience processing, response inhibition, interoception, and executive functions with neuroimaging can inform the development of pharmacological, neuromodulatory, and psychotherapeutic interventions to modulate the disordered neurobiology. Based on our systematic search, 409 protocols registered on ClinicalTrials.gov include the use of one or more neuroimaging paradigms as an outcome measure in addiction, with the majority (N=268) employing functional magnetic resonance imaging (fMRI), followed by positron emission tomography (PET) (N=71), electroencephalography (EEG) (N=50), structural magnetic resonance imaging (MRI) (N=35) and magnetic resonance spectroscopy (MRS) (N=35). Furthermore, in a PubMed systematic review, we identified 61 meta-analyses including 30 fMRI, 22 structural MRI, 8 EEG, 7 PET, and 3 MRS meta-analyses suggesting potential biomarkers in addictions. These studies can facilitate the development of a range of biomarkers that may prove useful in the arsenal of addiction treatments in the coming years. There is evidence that these markers of large-scale brain structure and activity may indicate vulnerability or separate disease subtypes, predict response to treatment, or provide objective measures of treatment response or recovery. Neuroimaging biomarkers can also suggest novel targets for interventions. Closed or open loop interventions can integrate these biomarkers with neuromodulation in real-time or offline to personalize stimulation parameters and deliver the precise intervention. This review provides an overview of neuroimaging modalities in addiction, potential neuroimaging biomarkers, and their physiologic and clinical relevance. Future directions and challenges in bringing these putative biomarkers from the bench to the bedside are also discussed.

## INTRODUCTION

Substance use disorders (SUDs), including alcohol, cause significant and increasing mortality and morbidity worldwide ^1,2^. In the United States alone, yearly costs of medical care, lost productivity and law enforcement associated with SUDs exceed an estimated $700 billion ^3^. As the designation suggests, SUDs have conventionally been viewed as disorders of “substance use” ^4^; but increasing evidence suggests that this harmful substance use is both driven by and contributes to pervasive brain alterations which underlie profound cognitive and behavioral manifestations broader than substance use ^5^. Since early pneumoencephalography studies revealed general brain atrophy in chronic alcohol users ^6^, decades of neuroimaging research have increasingly caused a shift towards a “brain disease” model of SUDs ^7–9^. Under this neuroimaging-informed model, genetic, developmental, social, and biological influences converge on combinations of core neurocognitive aberrations: the mesocorticolimbic reward network is sensitized by drugs of abuse, leading to excessive attribution of salience to drug-associated stimuli; anti-reward and stress systems across the basal ganglia and the extended amygdala become over-reactive, contributing to withdrawal symptoms and negative-affective states which can motivate substance use; and executive control networks centered around prefrontal regions are disrupted, with the degradation of top-down frontal control leading to disinhibited substance use ^7–11^.

Considering the evidence for neural aberrations in SUDs that can be objectively assessed using neuroimaging technologies, there is growing interest in using neuroimaging to inform clinical care and intervention development for SUDs ^12,13^. Objective measures of SUDs are currently limited to measures of psychoactive substances or their metabolites in biological samples (National Institutes of Health, 2020a) or reflect toxic effects of use^14^. These measures of substance use are not informed by the neurocognitive processes which underlie addiction, and thus have limited use in distinguishing at-risk individuals, offering prognostic insight, or informing interventions ^8^. In this context, neuroimaging technologies provide objective measures which could be used as novel “biomarkers” for SUDs, enabling the translation of neuroscientific insights to the bedside ^15^. This echoes broader trends in precision psychiatry and efforts to develop and utilize so-called “biomarkers” in psychiatric practice and research more extensively ^16,17^. Neuroimaging biomarkers, which can indicate specific aberrations of brain structure and function in SUDs, bring a three-fold advantage: first, they provide a direct window into proximal potential neurobiological mechanisms of disease and recovery in individuals with SUDs; second, they suggest novel treatment targets and provide neurophysiological evidence of effectiveness to facilitate intervention development; and third, mechanistically-grounded markers could be used directly for clinical purposes: to distinguish different sub-populations of substance-using individuals and inform personalized interventions and ongoing monitoring tailored for patients with specific brain abnormalities ^18–22^.

It is important to note that the “brain disease” model is not the only account of addiction etiology. For example, alternative explanations posit that addiction is a disease of choice and may be caused by a lack of alternative reinforcers ^23^, some contest whether addiction is a “disease” ^24^, and others simply argue that neurobiological explanations cannot be privileged over others ^25^. Moreover, the “brain disease” model has faced criticism on scientific, philosophical, and political grounds ^26–28^; and while it is generally agreed that alcohol and substance use disorders involve brain changes ^22,29^, some have argued that the current body of neurobiological evidence may not be sufficient to conclude that neurobiological dysfunctions are specific and primary causes of addiction broadly ^30^. However, while we would argue that the addiction neuroimaging literature to date both aligns with a “brain disease” model of addiction and supports the development of neuroimaging biomarkers, adherence to the former is not strictly necessary for the latter: According to the FDA-NIH Biomarker Working Group, a biomarker is simply “a defined characteristic that is measured as an indicator of normal biological processes, pathogenic processes, or biological responses to an exposure or intervention, including therapeutic interventions” ^31^. Regardless of whether addictive disorders are primarily caused or sustained by neurological dysfunction, neuroimaging biomarkers of aberrant brain structure or function associated with specific mechanisms of addiction and recovery could illuminate neural pathology, facilitate intervention development, and guide clinical care. A pertinent example is hypertension: the fact that the disease can be caused in large part by social and environmental factors does not diminish the importance of blood pressure as a biomarker to diagnose and monitor hypertension and develop interventions ^32,33^.

To lay the conceptual framework for a discussion of potential neuroimaging biomarkers in SUDs, we will provide an overview of the current status of neuroimaging paradigms in translational addiction neuroscience, informed by a systematic review of neuroimaging outcome measures in 409 protocols registered on ClinicalTrials.gov between its inception and November 17, 2021. Together, the 409 protocols have utilized 479 imaging modalities, and 688 neuroimaging outcome measures and provide a broad estimate of the clinically-relevant uses of neuroimaging in addiction neuroscience. We supplement this discussion with another systematic review of 61 meta-analyses between inception and November 10, 2023 of neuroimaging biomarkers in SUDs, and highlight biomarkers that have replicated in meta-analyses across multiple contexts and diagnoses. We then discuss different neuroimaging biomarkers which may be developed for SUDs based on taxonomy developed by the FDA-NIH Biomarker Working Group ^34^ and highlight challenges and future directions to provide clinicians and researchers with an understanding of opportunities and challenges in neuroimaging biomarker research.

## RESULTS

The present manuscript is informed by two systematic reviews. The first covered SUD clinical research protocols which include neuroimaging outcome measures, obtained by querying the ClinicalTrials.gov repository between inception and November 17, 2021 (Supplementary Figure 1a). This systematic review yielded a final result of 409 protocols. The second systematic review was conducted on PubMed, focusing on meta-analyses of neuroimaging studies of SUDs and finding 61 meta-analyses from which 83 meta-analytic findings were extracted (Supplementary Figure 1b). In this paper, while we seek to structure the discussion around replicated findings that have held across SUDs, some findings pertain only to specific SUDs, in which cases the particular SUD is highlighted. It should also be noted that the neuroimaging measures and findings in included protocols and meta-analyses do not constitute validated biomarkers: Any objective measure needs to undergo an extensive validation process to qualify as an actual biomarker of disease or recovery, which is not the case for any of the measures we discuss. Essential validation steps are discussed in the future directions. The systematic reviews serve to highlight replicated neuroimaging findings in SUDs and demonstrate the different exploratory purposes for which neuroimaging modalities are already used in clinical research. These purposes or “contexts of use” are grouped under corresponding categories of biomarkers to outline what measures might come to serve as actual neuroimaging biomarkers of SUDs, and motivate a discussion of challenges which need to be surmounted in the process.

## NEUROIMAGING MODALITIES IN ADDICTION MEDICINE

Interest in clinical uses of neuroimaging paradigms for virtually all SUDs has increased over time, with 87.3% of the protocols in our systematic review starting in 2010 or later. This is particularly the case with alcohol (N=139) and nicotine use disorders, but a growing number of protocols are using neuroimaging as an outcome measure for cocaine (N=44), cannabis (N=36), and opioid use disorder (N=31) (Figure 1a and Supplementray Figure 2). The growing interest in using neuroimaging paradigms has also been reflected in the conducted meta-analyses (Note that three of the studies are mega-analyses rather than meta-analyses, though we use the term meta-analysis to refer to these for simplicity). with all of them conducted after 2011 and more than half of them (N=31 out of 61) in the last 3 years. Most of the meta-analyses were conducted on multi substances (N=28) followed by analyzing studies focusing solely on alcohol (N=13) (Supplementary Figure 3). With some exceptions, neuroimaging paradigms in addiction neuroscience can be broadly categorized into “structural” imaging techniques which probe brain structure statically, “functional” paradigms which evaluate changes in a signal associated with brain function during the scan, and “molecular” paradigms which assess the static or changing distribution of important molecules/metabolites within the brain. These various paradigms are converging on a multi-scale perspective into brain changes in SUDs and may be used to develop clinically-relevant biomarkers ^35,36^.

**Figure 1:**
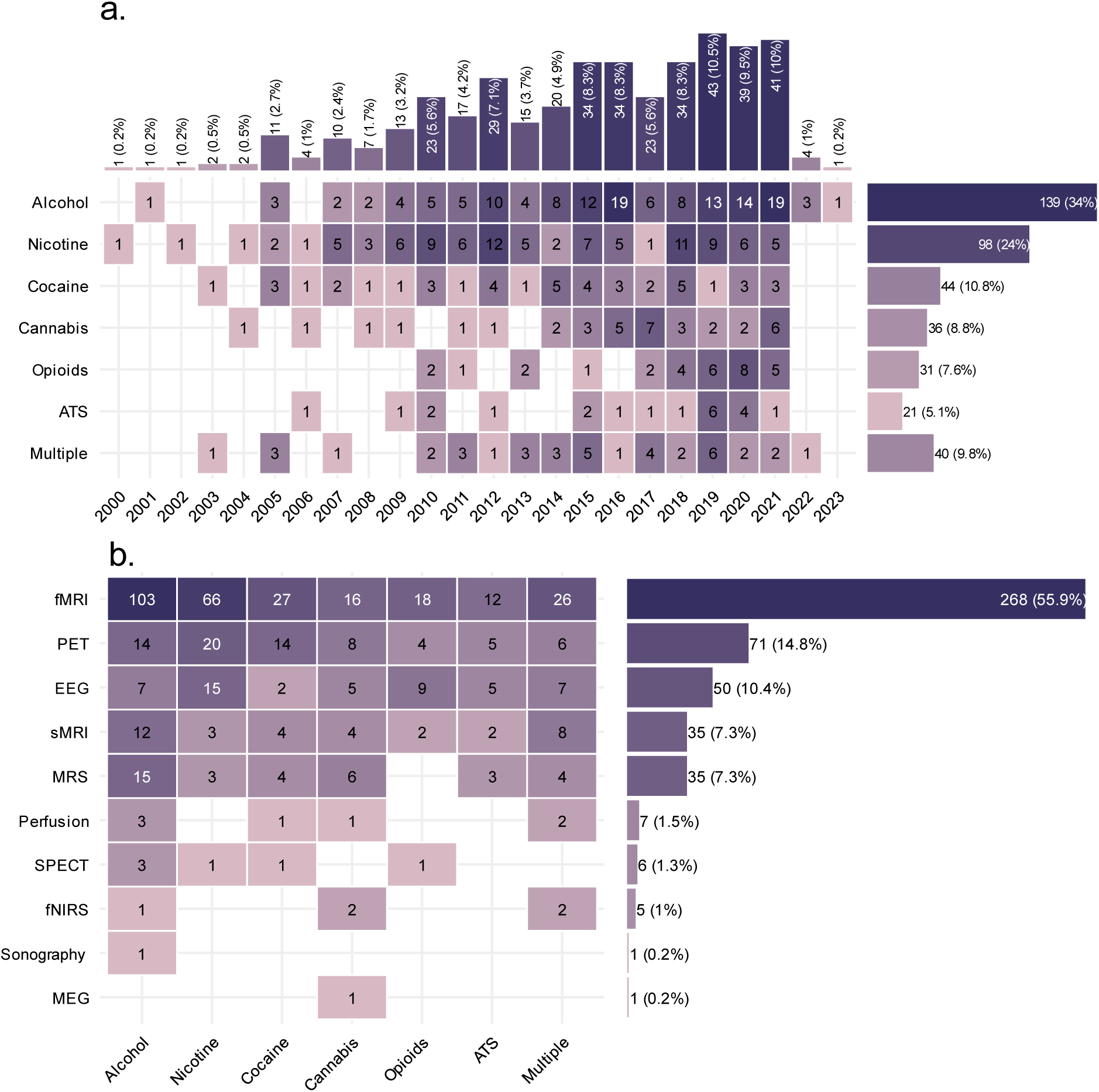
Distribution of the neuroimaging protocols based on year and substance. a. Number of protocols starting for each substance each year (n= 409). Years are obtained from the ClinicalTrials.gov database indicating actual or planned start years. b. Number of neuroimaging modalities used in each protocol for each substance. Numbers on this figure sum to 479 for 409 protocols, since 70 protocols used multiple imaging modalities. ATS: Amphetamine-type Stimulants; sMRI: structural MRI, including whole-brain T1 imaging, gray matter volumetry, or diffusion tensor imaging; Perfusion: brain perfusion imaging, including arterial spin labeling, cerebral blood flow imaging, and magnetic resonance angiography; MRS: magnetic resonance-spectroscopy. Data were collected from ClinicalTrials.gov on November 17, 2021.

**Figure 2:**
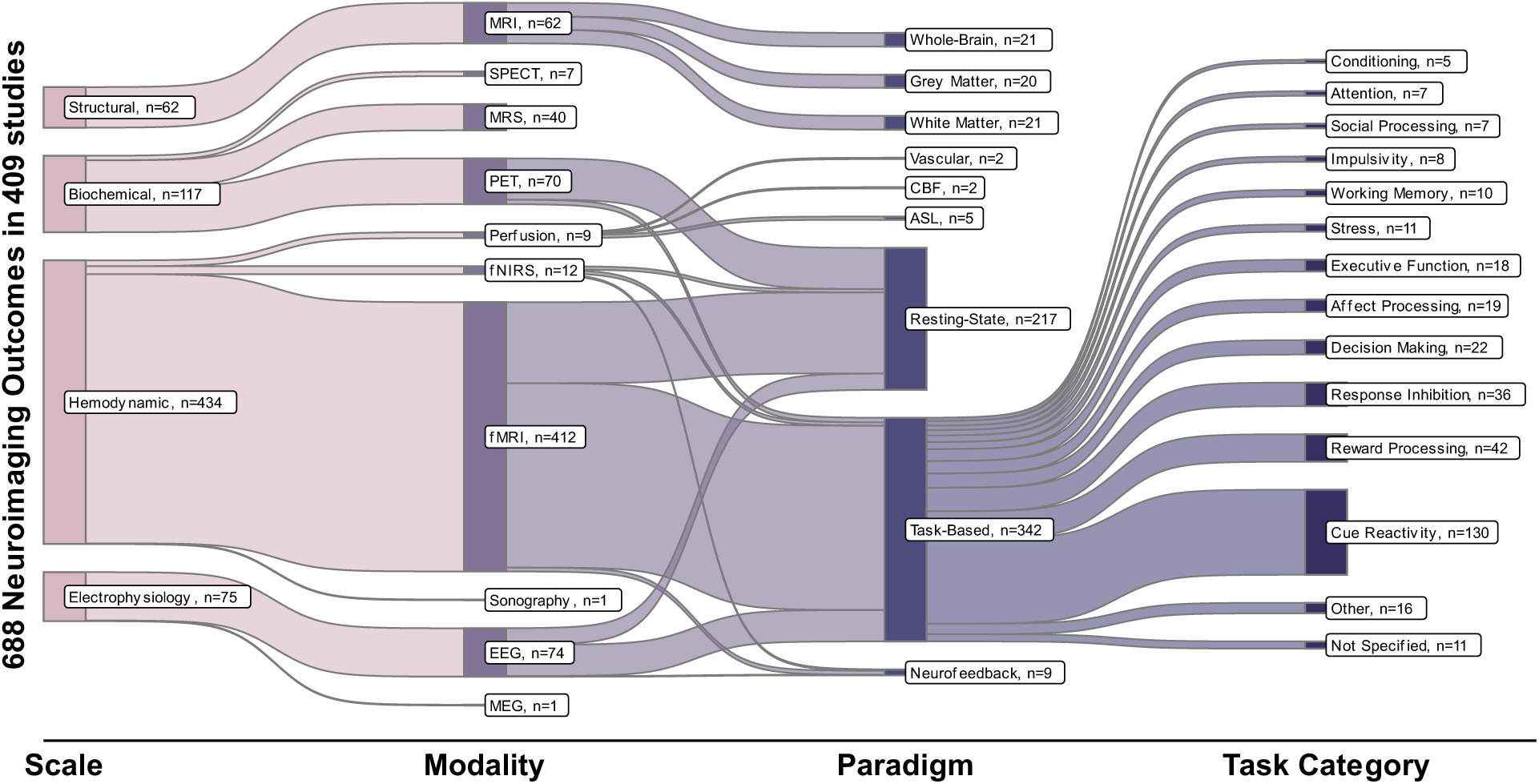
Multi-level characteristics of 688 neuroimaging outcome measures in 409 registered protocols. These levels include the scales at which neuroimaging modalities have probed the nervous system (structural, biochemical, hemodynamic or electrophysiology), the neuroimaging modality, different paradigms in each modality, and the types of tasks used in task-based functional neuroimaging paradigms. All “structural” paradigms in our database were variants of MRI; “biochemical” paradigms include SPECT, MRS, and PET; “hemodynamic” paradigms include fMRI, fNIRS, less common perfusion imaging modalities, and ultrasound; and EEG and MEG constitute “electrophysiological” imaging paradigms. These modalities have been used for static structural scans of brain gray or white matter and vasculature, resting-state functional scans, or task-related functional scans with various tasks. Note that many protocols have utilized more than one neuroimaging outcome measure and the total number of outcome measures is 688, more than the number of protocols (n=409). Data is collected from ClinicalTrials.gov on November 17, 2021. EEG: electroencephalography; *fMRI:functional* magnetic resonance imaging; fNIRS: Functional near-infrared spectroscopy; MEG: magnetoencephalography; *MRI:* magnetic resonance imaging; *MRS: magnetic resonance-spectroscopy; PET:* positron emission tomography; *SPECT: Single-photon emission computed tomography;*

### Brain Structure: Gray and White Matter

While a few studies have utilized CT scans to interrogate brain structure alterations in SUDs ^37^, arguably the most popular structural neuroimaging paradigm in addiction neuroscience is structural magnetic resonance imaging (sMRI), used by 35 protocols in our trials database as the only neuroimaging paradigm and by 27 protocols in conjunction with another paradigm (Figure 1b). Among the meta-analytic findings reviewed, 22 out of 83 were aberrations observed with structural MRI. Using MRI, algorithms such as voxel-based morphometry can isolate and quantify gray matter ^38^, and meta-analyses of these and similar techniques have revealed wide-spread losses of gray matter across cortical and subcortical regions across a number of different SUDs ^39–44^, though there is some evidence that these may recover with abstinence ^45^. “Mega-analyses” of MRI data collected from thousands of individuals with a variety of SUD types have also revealed an overall loss of gray matter, particularly in the insula and prefrontal and parietal cortices, and suggest that use severity may be correlated with lower amygdala and nucleus accumbens volume, particularly in alcohol use disorder ^46^. Simultaneously, studies of white matter structure with diffusion-weighted imaging have broadly revealed white matter degeneration in commissural tracts, the internal capsule, and corpus callosum across several SUDs ^39,47–49^. Observed structural changes in the gray and white matter might explain both deficits in higher-order cognitive processes and bottom-up processes in SUDs, with striking alterations in both frontal, parietal, and insular cortical regions involved in interoception, attention, and executive control and in the amygdala and nucleus accumbens which subtend bottom-up reward and affective processing ^46,50^.

### Brain Function: Hemodynamics and Electrophysiology

While structural neuroimaging paradigms are useful, the brain is engaged in constant activity during task performance and even idleness or sleep ^51^ and alterations in these rich neural dynamics underlie the cognitive-behavioral profiles typical of SUDs ^52^. This necessitates the use of “functional” neuroimaging paradigms that can measure brain activity either during the performance of various tasks (“task-based” imaging, 342 out of 688 instances in protocol database and 30 out of 83 in our meta-analysis database) or during rest (“resting-state” imaging, 217 instances in our protocol database and 4 in our meta-analysis database) ^53^. For example, “cue-reactivity paradigms” involve the presentation of stimuli associated with substances, such as pictures, scents or tastes, to assess neural reactivity and sensitization to these cues ^54^, and are used by 130 protocols in our protocol database (and 10 meta-analytic findings in SUDs). Other tasks can be used to probe other aspects of reward processing (42 instances across protocols, 6 meta-analytic findings), response inhibition (36 instances across protocols, 3 meta-analytic findings) and decision making (22 instances across protocols, 1 meta-analytic finding), all processes whose neural circuitry is impacted in SUDs ^11,55^ (Figure 2 and 3).

**Figure 3:**
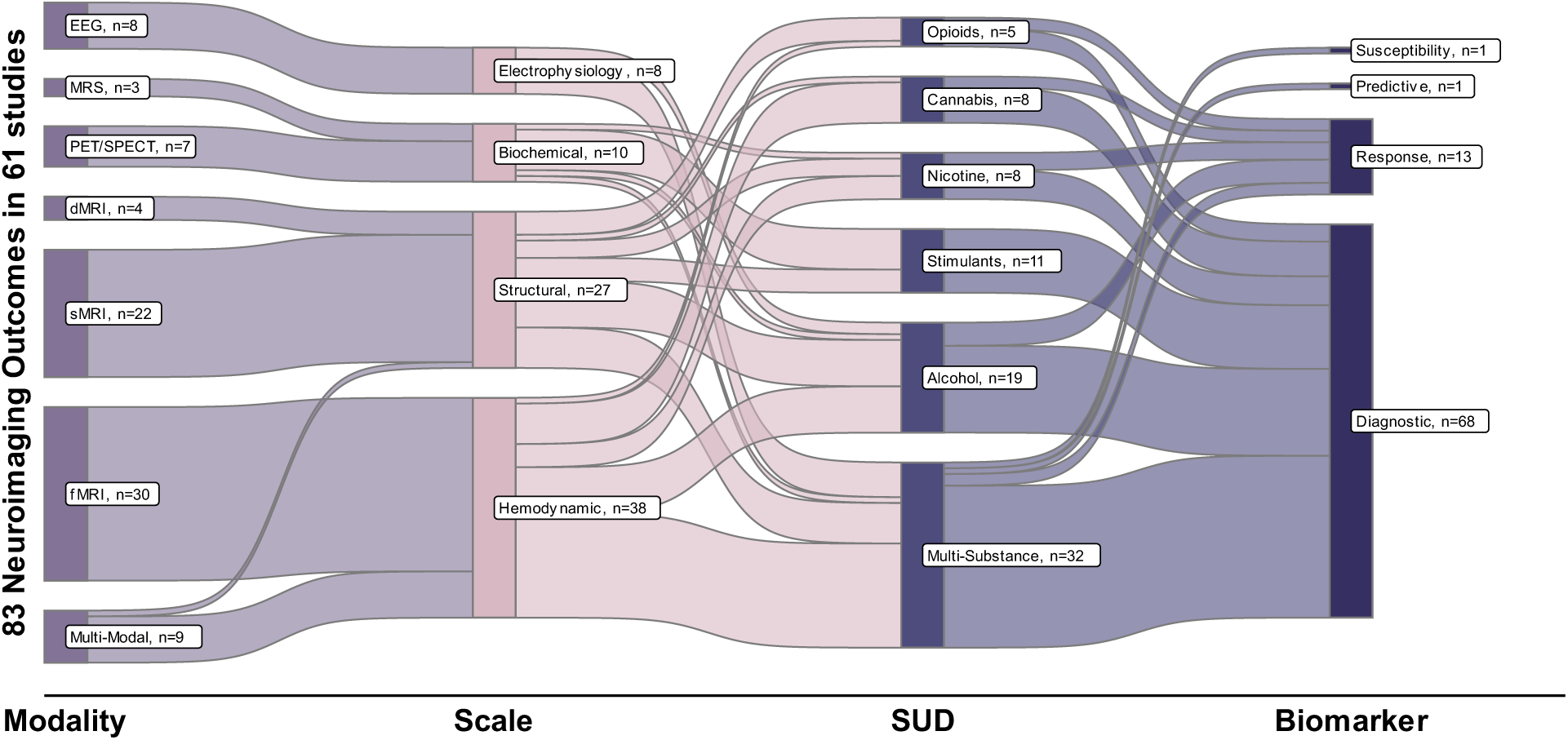
Multi-level characteristics of 83 neuroimaging outcome measures in 61 meta-analyses. These levels include the scales at which neuroimaging modalities have probed the nervous system (structural, biochemical, hemodynamic or electrophysiology), the neuroimaging modality, different paradigms in each modality, and the types of tasks used in task-based functional neuroimaging paradigms. All “structural” paradigms in our database were variants of MRI; “biochemical” paradigms include SPECT, MRS, and PET; “hemodynamic” paradigms include fMRI, fNIRS, less common perfusion imaging modalities, and ultrasound; and EEG and MEG constitute “electrophysiological” imaging paradigms. These modalities have been used for assessment of people with different kinds of SUDs. These assessment can be categorized into different biomarker categories. Note that some meta-analyses have utilized more than one neuroimaging outcome measure and the total number of outcome measures is 83, more than the number of total meta-analyses (n=61). Further, 3 of the 83 findings are from mega-analyses rather than meta-analyses, though we use the term meta-analysis to refer to these for simplicity. dMRI: diffusion magnetic resonance imaging; EEG: electroencephalography; *fMRI: functional* magnetic resonance imaging; fNIRS: Functional near-infrared spectroscopy; MEG: magnetoencephalography; *MRI:* magnetic resonance imaging; *MRS: magnetic resonance-spectroscopy; PET:* positron emission tomography; *sMRI: structural* magnetic resonance imaging; *SPECT: Single-photon emission computed tomography; SUD: Substance user disorder*

The first major group of functional neuroimaging outcomes (433 instances across protocols and 30 findings across meta-analyses) is “hemodynamic” techniques that include blood oxygenation level-dependent (BOLD) and arterial spin labeling (ASL) functional MRI, functional near-infrared spectroscopy, and cerebral perfusion imaging methods ^56–59^. fMRI is the most commonly used neuroimaging paradigm in addiction neuroscience, with 412 instances of fMRI as an outcome measure in our database out of the 688 neuroimaging outcome measures used in the 409 protocols (Figure 1b, Figure 2).

Further, 39 meta-analytic findings across neuroimaging SUD studies are from meta-analyses which include fMRI studies, alone (30 findings) or in combination with other modalities (Figure 3). There is extensive task-based fMRI evidence of disruption during reward-processing ^60^, and drug cue exposure results in a cascading hyperactivation of limbic circuits which subtend valuation and salience processing and disruption of prefrontal control, which can end in drug use ^55,61^. On the other hand, resting-state fMRI studies have revealed that SUDs are associated with weaker connections in the executive control network and stronger couplings within and between salience, reward, and “default mode” networks, suggesting that this might account for impaired response inhibition and the abnormal salience of drugs ^11,62^. Other hemodynamic paradigms have converged on similar findings, with aberrant function and perfusion in the middle frontal and orbitofrontal cortices, among others, observed in SUDs ^57,63^

The second group of functional imaging modalities focuses on the brain’s electrophysiological properties: electroencephalography (EEG) and magnetoencephalography (MEG) record, respectively, the electrical and magnetic fields generated during brain activity using extra-cranial probes to infer the underlying brain activity ^64,65^. Owing to its low cost and portability, EEG is the more common paradigm with 74 instances in our protocol database (and 8 findings in our meta-analysis database) compared to a single protocol with MEG. Event-related potentials elicited during task performance are usually split into components associated with underlying cognitive processes. For example, There is evidence that the P300 component of ERPs elicited by drug cues may be associated with reward valuation and the late positive potential with drug use motivation in individuals with SUDs, while the error-related negativity and feedback-related negativity components are associated with cognitive control and self-regulation ^66,67^. Another approach is to decompose the recorded EEG or MEG signal into specific “bands” with different frequencies, which has revealed decreases in EEG beta band power in opioid and alcohol use disorders ^68^. As with fMRI and fNIRS, EEG recordings also revealed network-level changes in individuals with SUDs: Examples include disruptions in the communication of the parietal lobe with other brain regions ^69^ and reductions in global integration and locally specialized connectivity ^70^.

### Brain Biochemistry: Molecular Systems

On a molecular level, positron emission tomography (PET) and single photon-emission computed tomography (SPECT) use radiotracers with specific patterns of distribution across the tissue. Psychiatric SPECT and PET imaging increasingly use complex ligands known to preferentially bind to molecules of interest to probe both the density and binding potential of a certain neurotransmitter system across the brain, and dynamic changes in neurotransmission induced by a pharmacological agent or during cognitive and behavioral tasks ^71^. Magnetic resonance spectroscopy (MRS) is a different approach to investigating molecular concentrations across the brain, using magnetic resonance rather than ionizing radiation to assess relative levels of different metabolites, such as choline and N-acetylaspartate, and neurotransmitters, such as glutamate, GABA, and glutamine ^72^.

All three modalities are used in our protocol database as outcome measures, with 70 instances of PET, 7 instances of SPECT, and 40 instances of MRS (Figure 2), and 16 meta-analytic PET/SPECT findings (7 findings from meta-analyses of studies using only PET/SPECT, 9 in combination with fMRI studies). PET and SPECT studies have demonstrated that dopamine transporter and D2 dopamine receptor availability are consistently downregulated in SUDs, especially D2 receptors in the striatum whose downregulation is associated with compulsive drug use ^73^. This has been extensively corroborated in stimulant use disorders, with several recent meta-analyses ^74,75^. These observations and further aberrations in dopamine synthesis and release are consistent with dysfunctional dopaminergic neuroadaptations in the reward network and accompany changes in other neurotransmitter systems implicated in the neuro-cognitive abnormalities observed in SUDs, such as serotonergic disruptions potentially related to affective deregulation and opioidergic down-regulation which may explain tolerance and dependence ^76–78^. At the same time, meta-analyses of MRS studies have revealed decreased N-acetylaspartate levels across frontal and cingulate regions, suggesting decreased neuronal and axonal viability ^79,80^; and others have reported aberrations in glutamate and GABA levels in the prefrontal cortex and basal ganglia which correlate with disease severity and cognitive function across SUDS ^81,82^. These findings suggest that neurotransmitter abnormalities may account for some neuro-cognitive abnormalities in attention and executive function observed in SUDs.

## NEUROIMAGING BIOMARKERS IN ADDICTION

Given the observation of brain abnormalities across different domains in SUDs, there are ongoing efforts to utilize these brain aberrations as biomarkers for specific contexts of use. The neuroimaging technologies discussed above have distinct advantages and disadvantages, and thus each may be better suited for use in certain contexts and/or for different SUDs. The systematic review of the registered protocols discussed above expectedly identified mostly neuroimaging biomarkers used to measure the effect of an intervention in a trial. However, neuroimaging biomarkers could go beyond treatment response assessment. The FDA-NIH Biomarker Working Group has formally defined distinct biomarker types which correspond to different stages of addiction, recovery, and clinical intervention ^34^: In the context of SUDs, “susceptibility” biomarkers indicate the risk that individuals develop a SUD and “diagnostic” biomarkers can distinguish individuals with SUDs from recreational users or between clinically relevant subtypes of SUDs. For individuals with an established SUD diagnosis, “prognostic” biomarkers can predict the future progression of patients towards relapse versus remission and “monitoring” biomarkers can be measured over time to assess changes.

When developing or implementing a clinical intervention for SUDs, “predictive” biomarkers can predict the clinical impact of an intervention, and “safety” biomarkers can be measured to assess the safety of an intervention or novel substance; while “response” biomarkers reflect an individual’s response to an intervention and, under certain conditions, can be used as “surrogate endpoints”: biomarkers which can demonstrate the likely clinical effectiveness of an intervention before actual clinical outcomes develop ^83,84^. A schematic of the different stages of SUDs and intervention is presented in Figure 4. It is important to note that a single neuroimaging measure may conceivably serve multiple biomarker roles in different contexts: as an example, higher baseline ventral striatal fMRI drug cue-reactivity can distinguish relapsing individuals with stimulant use disorder from non-relapsing individuals 3 months after the scan (prognosis) ^85^ and predict the clinical response of individuals with alcohol use disorder to naltrexone (prediction) ^86^. At the same time, striatal cue-reactivity in individuals with alcohol use disorder can be reduced through treatment (response) ^87^. Such converging evidence can support the clinical validity of a biomarker.

**Figure 4:**
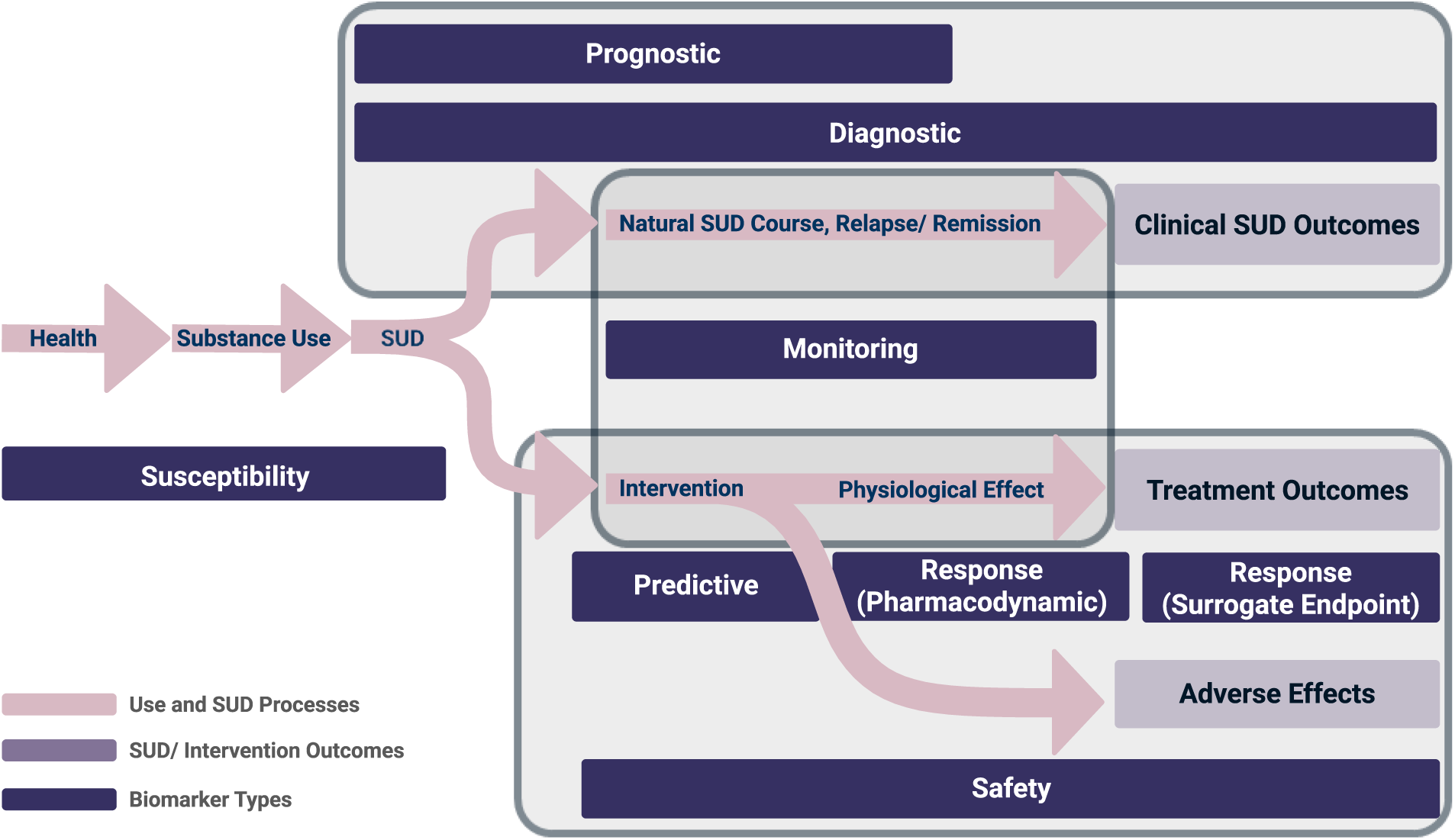
Schematic representation of stages in substance use and SUDs and their therapeutic interventions and corresponding biomarker types. Susceptibility biomarkers can predict transition to substance use or disorder, prognostic biomarkers can predict the future progression of the disorder, diagnostic biomarkers can distinguish clinically-relevant populations, monitoring biomarkers facilitate ongoing information about the course of the disorder with or without intervention, predictive biomarkers can predict treatment response, response biomarkers can reflect the physiological impact of an intervention and potentially be used as surrogate endpoints in lieu of clinical outcomes, and safety biomarkers can help assess the potential hazards of various substances used in clinical or non-clinical settings.

Based on an assessment of the structure of the reviewed protocols, the 409 protocols have collectively used 510 neuroimaging measures as putative SUD biomarkers. These 510 putative neuroimaging-based biomarkers are broken down based on biomarker type, substances, and neuroimaging modalities in Figure 5. Based on the systematic review of meta-analyses, several of these markers have also been suggested across several SUDs or contexts of use in meta-analyses of neuroimaging studies. Such suggested findings were observed in 55 meta-analyses in our database and are summarized in Table 1. The following sections review these biomarker types in greater detail.

**Figure 5:**
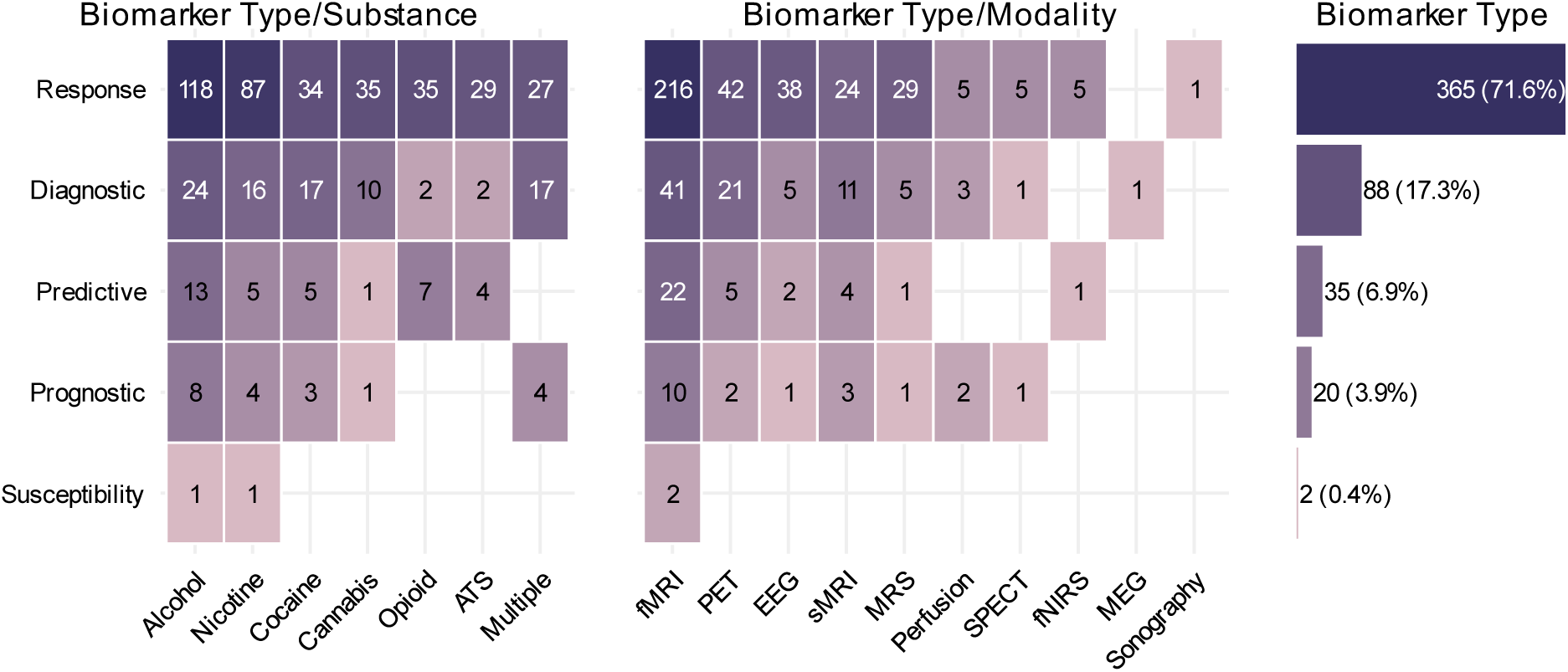
Putative neuroimaging biomarkers reported in registered protocols in various substance use disorders (SUDs) and neuroimaging modalities. Biomarker types are divided between the substance of interest and neuroimaging modalities used in the protocol (510 biomarkers across 409 protocols). The horizontally aligned bars represent the total number of each biomarker type. Note that some of the protocols include more than one biomarker type. Some protocols did not report enough details for neuroimaging modalities in a way that fit any biomarker’s definition. Data is collected from ClinicalTrials.gov on November 17, 2021.

**Table 1:** Meta-analytic neuroimaging markers that have been suggested across SUDs or contexts of use. Note that this table includes only findings supported by more than one meta-analyses across SUDs or contexts of use, and thus only 55 out of the 61 meta-analyses in the full database are included. *COU: Context of Use* dMRI: diffusion magnetic resonance imaging; EEG: electroencephalography; *fMRI: functional* magnetic resonance imaging; fNIRS: Functional near-infrared spectroscopy; MEG: magnetoencephalography; MRI: magnetic resonance imaging; *MRS: magnetic resonance-spectroscopy; PET:* positron emission tomography; *sMRI: structural* magnetic resonance imaging; *SPECT: Single-photon emission computed tomography; SUD: Substance user disorder*

| Modality | Number of Meta-Analyses | SUD | COU | Observations | References |
| --- | --- | --- | --- | --- | --- |
| **Gray and White Matter** |  |  |  |  |  |
| dMRI<br>(last study White Matter VBM) | 6 | Alcohol,<br>Stimulants,<br>Opioids | Diagnostic,<br>Response | Macro- and microstructural evidence of white matter degeneration across the corpus callosum, internal capsule, and frontal and limbic projections. Evidence of white matter recovery with abstinence at least in alcohol use disorder | 39,47–49,221,222 |
| sMRI | 18 | Alcohol, Nicotine,<br>Stimulants,<br>Opioids,<br>Cannabis | Diagnostic,<br>Response | Reduction in cortical thickness and gray matter volume across superior temporal, inferior parietal, precentral, insular, frontal, cingulate, hippocampal and parahippocampal cortices and the striatum and thalamus. Further, at least in the case of nicotine, agnosts impact some of the brain areas where reductions in gray matter volume are prominent. | 39–45,132,133,194,212,223–229 |
| **Neurotransmitter Systems and Metabolites** |  |  |  |  |  |
| PET/SPECT | 2 | Stimulants | Diagnostic | Overall downregulation of striatal dopaminergic signaling, including decreases in dopamine release, reduced dopamine transporter density and availability, and reduced dopamine receptor density, availability and binding potential. | 74,75 |
| MRS | 2 | Alcohol,<br>stimulants | Diagnostic | Lower N-acetylaspartate levels across frontal and cingulate regions suggesting decreased neuronal and axonal viability, lower cortical and higher subcortical creatine levels. | 79,80 |
| **Electrophysiological Activity** |  |  |  |  |  |
| EEG (Response Inhibition) | 3 | Alcohol, General (Opioids, Stimulants, Nicotine, Cannabis) | Diagnostic, Response | SUDs are associated with the attenuation of error-related negativity and EEG components such as N200. Alcohol administration leads to acute reduction of ERN. | 198,206,230 |
| EEG (Cue-Reactivity) | 2 | General (stimulants, opioids, alcohol, nicotine) | Diagnostic, Response | SUDs are associated with the enhancement of the salience-related P300 potential in response to drug-related cues, which also shows signs of time-dependent recovery with abstinence. | 197,198 |
| EEG (Attention and Surprise) | 2 | General (alcohol, opioids, nicotine, stimulants, others) | Diagnostic, , Susceptibility | Reduced P300 amplitude in response to tasks which involve attention and surprise (such as the oddball paradigm) is associated with SUDs, and may susceptibility to SUDs. | 204,231 |
| **Hemodynamic Activity** |  |  |  |  |  |
| fMRI/PET (Cue-Reactivity) | 11 | Alcohol, Nicotine, Stimulants, Opioids, Cannabis, General | Diagnostic, Susceptibility, Response | SUDs is associated with higher fMRI drug cue reactivity (FDCR) across mesocorticolimbic and nigrostriatal regions, the precuneus, cingulate and insula, various frontal and temporal regions, sensory cortices, and the cerebellum. FDCR may indicate susceptibility as well, particularly striatal FDCR in adolescents. Abstinence may lead to short-term hyperactivations in some of the regions, but in the long term treatment can normalize FDCR across regions, particularly striatum, insula, and prefrontal regions. | 96,135,137,195,199,200,232–236 |
| fMRI (Reward Processing) | 4 | Alcohol, General (alcohol, nicotine, stimulants, cannabis) | Diagnostic | Both anticipation and receipt of reward and loss are associated with pervasive hypo-and hyper-activations across striatal, prefrontal, orbitofrontal, sensory, insular and temporal cortices. | 60,136,138,237 |
| fMRI (Response Inhibition) | 3 | Alcohol, General (stimulant, alcohol, nicotine, opioid) | Diagnostic, | In SUDs compared to HCs, response inhibition is associated with lower activations across cingulate, frontal, inferior parietal, insular and temporal cortices. | 207–209 |
| fMRI (Rest) | 2 | General (stimulants, heroin, alcohol, cannabis, nicotine) | Diagnostic | Aberrant resting-state functional connectivity patterns across limbic, salience, frontoparietal and default-mode networks. | 238,239 |

### Neuroimaging Biomarkers for Assessment

The most straightforward application of neuroimaging biomarkers for SUDs would be for assessment purposes, since any structural, functional, or biochemical brain differences between individuals with and without SUDs could, hypothetically, be used to at least support the existence of disease. Accordingly. we identified 110 putative assessment neuroimaging markers in our systematic review of clinical research protocols and 69 across meta-analyses of neuroimaging studies in SUDs. However, mere diagnosis may not be the best use of neuroimaging biomarkers. Currently, diagnoses rely ultimately on relatively inexpensive clinical interviews, but these diagnostic criteria lead to heterogeneous patient populations, and moreover diagnoses rely ultimately on relatively inexpensive clinical interviews and the added benefit of biomarkers is unclear ^88^. More promising may be the use of neuroimaging biomarkers for clinically-relevant subtyping, prognosis, and patient monitoring.

Biomarkers for diagnosis, subtyping, and susceptibility assessment: Conceivably useful assessment neuroimaging biomarkers for SUDs fall into a few contexts of use. One would be “**diagnostic**” biomarkers which differentiate healthy and disordered substance use rather than individuals with SUD and non-drug users, given that distinguishing dependent and recreational use purely on the basis of self-report and drug use quantity is difficult ^89^. We identified 88 instances of potential diagnostic biomarkers across protocols (Figure 5) and 68 across meta-analyses (Figure 3) in our systematic review databases. Several neuroimaging biomarkers may help distinguish dependent and non-dependent users. For example, dependent compared to light alcohol use may be associated with greater alcohol-cue-induced BOLD signal in the dorsal striatum but lower signal in the ventral striatum ^90^ and dependent cannabis users have lower OFC volume compared to recreational users ^91^. Such diagnostic biomarkers may be especially relevant in the staging of SUDs, given the recently proposed category of “pre-addiction” ^92^. Another use of diagnostic biomarkers could be to distinguish SUD patients with the same diagnosis, but different underlying neuro-cognitive pathology. For example, heavy alcohol drinkers who drink primarily for “relief” from negative affect have greater alcohol-cue-induced BOLD signal in the dorsal striatum as compared to “reward” drinkers ^93^.

Another useful class of assessment biomarkers would be markers of “**susceptibility**”, biomarkers which predict the development of SUD in at-risk individuals in the absence of diagnosable disease. Only two of the registered protocols and one meta-analysis had putative susceptibility biomarkers, which require studying healthy participants for the development of SUDs. Much of the previous SUD-susceptibility neuroimaging research has been conducted in adolescents, who are particularly at risk of initiating substance use and transitioning to SUDs due to reward deficits associated with the striatal dopaminergic reorganization and the faster development of limbic emotion and reward systems compared to the prefrontal control circuitry ^94^. Consistent with this theory, task-related fMRI investigations have shown that dorsal striatal hyper-activation during reward tasks may be a marker of substance use vulnerability and is linked with co-existing externalizing psychopathology, and stronger responses of the reward-related nucleus accumbens and orbitofrontal regions to alcohol cues can distinguish individuals who transition to heavy drinking ^95,96^. Moreover, response inhibition fMRI studies have shown that blunted frontoparietal activity during inhibition and hyperactivation during inhibition failures predict the initiation of substance use ^97^. Structural MRI studies have converged on similar findings: Both lower volumes and lower white matter integrity in fronto-limbic regions involved in reward processing and decision-making may be markers of susceptibility to substance use initiation and the development of SUDs ^98^.

#### Biomarkers for prognosis and monitoring

With the rising number and larger sample sizes of studies with prospective and longitudinal designs, it has become possible to investigate relationships between neuroimaging parameters and subsequent clinical trajectories, enabling the development of “**prognostic**” biomarkers, with 20 examples in our systematic review of study protocols. An important clinical use of these biomarkers would be to predict relapse in abstinent individuals more accurately than is possible using self-report or behavioral task performance alone. Task-based fMRI studies have shown that individuals who require high neural activation for response inhibition are more prone to relapse, even with normative behavioral task performance ^97^, and baseline nucleus accumbens drug cue-reactivity may predict relapse with an accuracy outperforming conventional measures ^85^. Resting-state fMRI has further demonstrated that the weaker inter-regional synchrony in the executive control network may account for poorer response inhibition and can predict relapse ^99^.

Neuroimaging biomarkers that are measured over time can also be used as “**monitoring**” biomarkers, offering insights into the development and abatement of neuro-cognitive pathology to complement the clinical picture. These biomarkers are difficult to develop since they require repeated neuroimaging measurements and a model of their correspondence with clinical states and clinically-relevant phenotypes over time. None of the protocols or meta-analyses in our databases had the requisite structure to contribute to the development of potential monitoring biomarkers. Nevertheless, much of the research on using neuroimaging outcomes for putative monitoring markers has focused on neurological recovery during abstinence: longitudinal studies have shown that both gray and white matter degeneration in the frontal cortices of individuals with SUD can recover after abstinence ^100,101^, and in PET and SPECT studies striatal dopamine transporters downregulated in methamphetamine use disorder can recover during abstinence ^102,103^. A striking finding is the observation that individuals with SUD experience an “incubation” and accumulation of drug craving following abstinence which may predispose them to relapse. Another study has revealed that this “craving incubation” is reflected in the amplitude of the late positive potential, a marker of attention bias to drug cues which follows an expected parabolic trajectory during abstinence and a feature that would be missed by relying purely on self-report measures ^67^.

### Biomarkers for Intervention

Perhaps even more important than diagnostic, prognostic or susceptibility assessment of SUDs would be the use of neuroimaging biomarkers in interventional contexts; for example, to develop or implement interventions, objectively assess their neurophysiological impact in clinical trials or psychiatric practice or predict their outcomes and therefore serve to guide intervention selection. Furthermore, neuroimaging biomarkers of cognitive processes such as cue-induced craving and reward processing can directly become targets for intervention. According to our systematic review of ClinicalTrials.gov protocols, several multi-scale brain aberrations identified in observational studies of SUDs are under investigation as putative interventional biomarkers. Some of these are illustrated in Figure 6. Protocols with potential interventional biomarkers constitute a majority of the protocol database and contain 400 putative biomarkers. this is unsurprising since we reviewed ClinicalTrials.gov protocols, which mostly consist of interventional studies. Across meta-analyses however, there were only 14 examples of findings relevant to interventional contexts of use.

**Figure 6:**
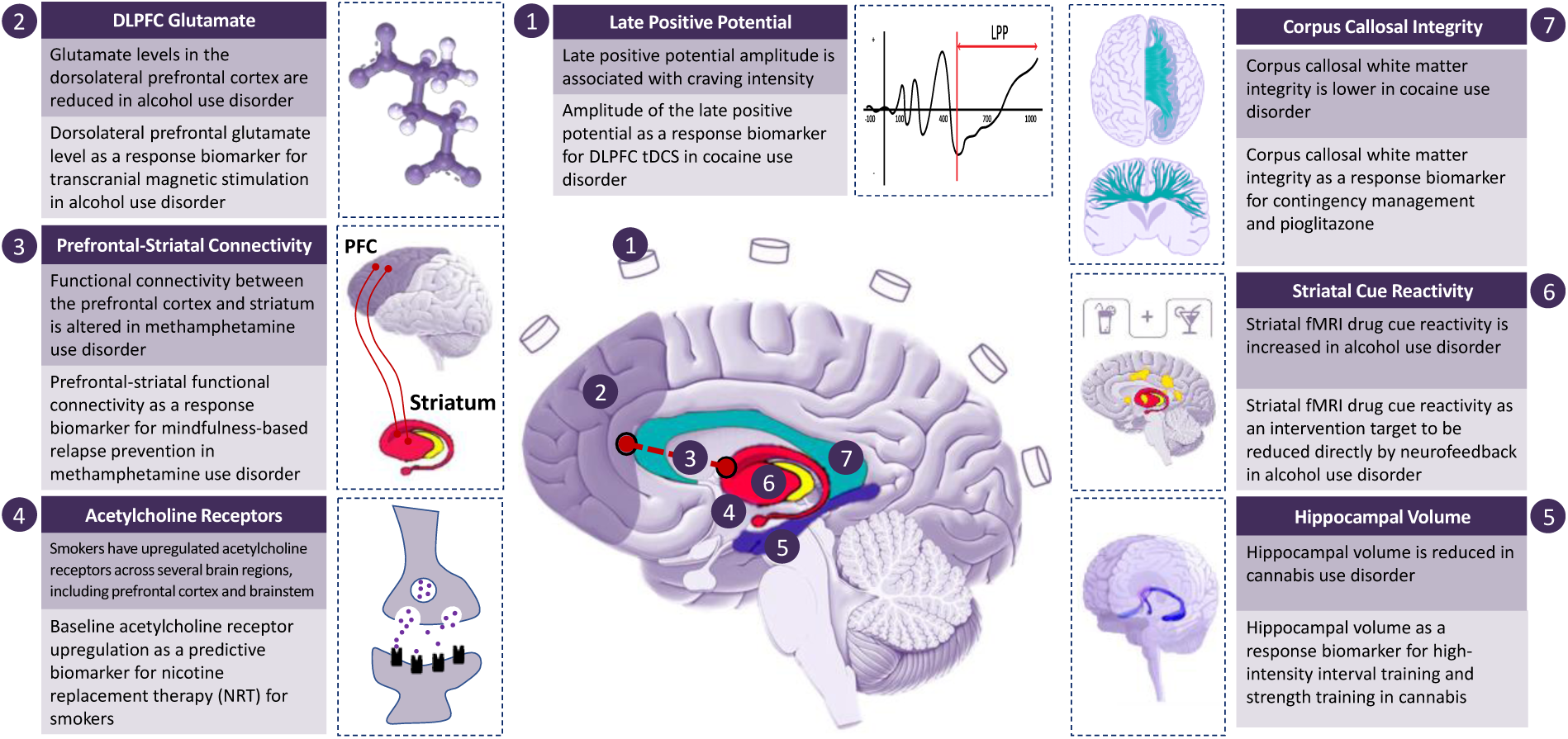
Multi-scale brain aberrations as putative neuroimaging biomarkers in trials for substance use disorders (SUDs) Seven examples of brain aberrations identified in SUDs (yellow boxes) that have been investigated as putative “response” or “predictive” biomarkers or intervention targets in protocols registered in ClinicalTrials.gov (light blue boxes). The relevant literature is referenced in supplementary table 2. FC: Functional Connectivity; FDCR: fMRI Drug Cue Reactivity; PFC: Prefrontal Cortex. tDCS: Transcranial Direct Current Stimulation.

#### Biomarkers of intervention response and safety

The effectiveness of interventions for SUDs is generally assessed by measuring their impact on drug use, which provides little information about neuro-cognitive recovery ^8^. A paradigmatic group of interventional biomarkers is “**response**” biomarkers. In early phases of intervention development, “**pharmacodynamic**” response biomarkers can indicate the presence of a treatment effect on neuroimaging biomarkers of recognized importance in SUDs and provide some estimate of the intensity and location of this effect. In our systematic reviews, 365 neuroimaging outcomes were used as putative response/pharmacodynamic biomarkers across protocols and 13 neuroimaging response markers were discovered across meta-analyses. Response biomarkers can be used to screen candidate therapeutics and prioritize those with plausible effectiveness, as in the “Fast-Fail” initiative of the National Institute of Mental Health^104^. In this context, research could be focused on therapies that engage brain substrates of SUDs. For example, pharmaco-fMRI studies have shown that baclofen can dampen increased drug cue-reactivity ^105,106^, and PET imaging can directly measure the dose-dependent impact of various therapies on neurotransmitter systems ^107^.

A narrower and more impactful subclass of “**response**” biomarkers are “**surrogate endpoints**”. These neuroimaging measures would not only correlate with the clinical effect of a therapy but causally lie along the physiological route between an intervention and its clinical effect in SUDs. A paradigmatic example of a surrogate endpoint in medicine is blood pressure, widely accepted as an outcome measure in clinical trials since it is known that anti-hypertensive medications offer clinical benefit through lowering high blood pressure, even though blood pressure in itself is not a clinical endpoint ^108^. Rigorous clinical trials might be able to establish that the impact of therapies such as dlPFC stimulation on craving is mediated through the modulation of cue-related neural activation and connectivity, leading to the development of surrogate endpoints ^63^.

Biomarkers assessed over time can be used as “**monitoring**” biomarkers in the context of interventions as well, establishing links between a neuroimaging biomarker and clinical response. For example, multiple imaging rounds in a trial of naltrexone for alcohol use disorder showed that naltrexone lowers ventral striatal fMRI drug cue-reactivity from baseline and greater reduction is associated with a larger clinical response ^109^, and event-related potentials recorded with EEG or MEG can be assessed during and after treatment to demonstrate the normalization of ERP components associated with attention bias or error-processing ^66^.

While we classified markers which show the neural impact of novel compounds as “response” biomarkers since their protocols did not explicitly use them to indicate the safety of interventions, neuroimaging biomarkers could also be used to gauge the **safety** and toxicity of various compounds of interest in addiction medicine. One example would be the use of neuroimaging to inform ongoing discussions on the safety of electronic cigarette products, where fMRI has been used to demonstrate that e-cigarette smoking may immediately induce activation across sensorimotor areas ^110^ and sweet-tasting products may synergize with nicotine content to increase the influence of e-cigarettes on nucleus accumbens reactivity ^111^. Another pertinent use-case is assessing the abuse potential of analgesic medications. Many such therapeutics, and in particular opioid medications, may lead to addictive substance use in some individuals, and neuroimaging biomarkers of safety may serve as early warning signs both during drug development and treatment ^112^. Neuroimaging safety biomarkers may also be useful to assess the brain impact of alcohol and opioid medications in individuals with genetic susceptibility to addiction, such as those with certain variants of dopamine and opioid receptor genes^113,114^.

#### Biomarkers for treatment targeting and implementation

Data on the effectiveness of current interventions for SUDs remains inconsistent, necessitating the development of more consistently efficacious interventions and subtyping individuals with SUDs to develop personalized treatment protocols/plans ^115^. Beyond providing information about the neural impact of treatment, neuroimaging biomarkers could enable individually targeted SUD treatment by reflecting a patient’s baseline or dynamically changing neural state. An example of this is targeting brain stimulation at important hubs of aberrant networks in each patient since electric and magnetic neuromodulation have connectivity-dependent effects (Siddiqi et al., 2019; Weigand et al., 2018) and it has been proposed that both structural and functional MRI can be used to optimally target the stimulation the inhibitory frontoparietal network in patients with SUD ^116^. The importance of targeting specific networks for intervention is further supported by recent observational evidence that brain lesions which affect areas functionally connected to cingulate, prefrontal, insular, and temporal regions can consistently induce remission in individuals with SUD ^117^. In addition to using baseline neuroimaging, more sophisticated technologies are paving the way for concurrent neuromodulation and brain imaging. These include TMS or tDCS with simultaneous EEG, MEG, fNIRS, or fMRI ^118–122^. These methods provide immediate readouts of the effects of neuromodulation on network activity and can be used to develop “closed-loop” stimulation systems where neuromodulation is dynamically adjusted for optimal impact^123^. Lastly, EEG and fMRI biomarkers that are correlated with undesirable SUD-related symptoms such as craving have been successfully used in neurofeedback training, where patients with tobacco or alcohol use disorder learned to attenuate these signals based on dynamic feedback ^124,125^.

#### Biomarkers to predict treatment effect

The final potential use case of biomarkers in an interventional context would be to **predict** the impact of therapies. We identified 35 neuroimaging outcome measures in our systematic review of protocols that serve as putative predictive biomarkers, though only one relevant marker was identified in the systematic review of meta-analyses. As the variability in the effectiveness of interventions for SUDs may be, in part, due to distinct baseline neurocognitive states, neuroimaging biomarkers could help the selection of interventions most likely to ameliorate the underlying pathology in each patient ^20,126^. For example, among individuals with AUD, a reduction of fMRI drug cue-reactivity in both the left putamen and the right ventral striatum can predict the effectiveness of naltrexone ^109,127^; for individuals with cocaine use disorder, greater persistence of the cue-triggered brain response across the cue task predicts poor drug use outcome ^128^. Machine-learning algorithms using task-related and resting-state fMRI data have been able to predict treatment response and completion in individuals with stimulant and heroin use disorders ^129,130^. Structural connectivity biomarkers may also have predictive value: reduced structural connectivity between the right anterior insula and nucleus accumbens at baseline can predict relapse to stimulant use up to six months after residential treatment ^131^.

Arguably, a robust neuroimaging biomarker of SUDs would be valid in several different contexts of use. Further, if the biomarker reflects physiological changes which are broadly important in the etiogenesis of SUDs and in recovery, such physiological changes would likely be detectable with different neuroimaging modalities and in different substance use disorders. Several neuroimaging markers with converging supporting evidence across meta-analyses have been discussed in Table 1, but a particularly promising set of examples are those which reflect the structure, function and connections of the striatum. Textbox 1 is dedicated to a discussion of findings of striatal involvement across SUDs, evidence supporting the use of striatal markers across neuroimaging modalities and contexts of use.

##### Textbox 1

**Striatal neuroimaging biomarkers in SUDs.** A discussion of the potential of neuroimaging markers of striatal function and structure across different contexts of use in SUDs and important next steps.

There is overwhelming evidence that the striatum is involved in the pathogenesis of SUDs. Meta-analyses have shown striatal atrophy across substance use disorders ^132,133^, impaired dopamine neurotransmission ^74,75^, and striatal dysfunction across substances and task paradigms, particularly in reward-related tasks and those which induce craving ^60,134–138^. Based on these observations and studies in animal models, major neuroscientific theories of addiction feature striatal dysfunction as a central cause of the aberrant reward processing, impulsivity, and incentive sensitization which drive _SUDs 7,139._

This body of literature, paired with relevant findings across contexts of use, provides an extensive foundation to support the clinical validation of neuroimaging biomarkers of striatal structure and function. As an example, striatal fMRI drug cue reactivity might indicate individual susceptibility to alcohol use disorder ^140^, diagnostically ^93^ and prognostically ^85^ demarcate clinically relevant subtypes of disease, predict treatment response ^86^, and reflect treatment response ^141^ *or monitor it across time* ^109^. An important next step would be investigating analyaitcal properties of striatal neuroimaging biomarkers, data on which is sparse. There is evidence supporting the longitudinal stability of striatal fMRI drug cue reactivity ^142,143^. There is also evidence for reasonable test-retest reliability of striatal PET imaging ^144,145^ and morphometry and cortico-straiatal integrity measures ^146^ in non-SUD samples; but these should be further replicated across larger samples with different SUDs.

Further, there is little formal guidance and consensus on best methodological practices for striatal neuroimaging, which may differ from those for cortical neuroimaging. For example, a 32-channel receiving coil may be more sensitive to cortical signals than an 8-channel coil but less sensitive to subcortical activations ^147^, and fMRI with higher field strengths seems to be more crucial for imaging the striatum than the cortex ^148^. Any striatal neuroimaging biomarker would need to be precisely *specified*, with methodological parameters, the target population, and standard operating procedures selected with respect to its context of use. This is since measures of striatal structure, function and connections are impacted by image acquisition parameters ^149^ and processing and reconstruction pipelines ^150^, behavioral task design ^151^, operating parameters such as time of day ^152^, and participant characteristics such as sex ^153^ and psychiatric comorbidity ^107^. Further research is required to clarify how these factors impact the clinical validity and analytical properties of striatal markers in different contexts of use and guide biomarker specification.

Lastly, a putative striatal biomarker needs to be cost-effective, but there has been virtually no cost-benefit analysis of any striatal neuroimaging biomarker. While most of the cited literature supporting the clinical use of striatal neuroimaging in SUDs has used functional neuroimaging paradigms, it is difficult to assess striatal function with

## CHALLENGES AND FUTURE DIRECTIONS

Despite decades of research highlighting the potential of neuroimaging technologies for the development and validation of biomarkers of SUDs and the proposal of several promising biomarkers in recent years ^13,154,155^, critics have noted that substantial investment in biomedical addiction research has not yet led to the development of biomarkers with substantial clinical utility ^156^. There is growing awareness of the myriad challenges ahead of pushing neuroimaging biomarkers through the “translational gap” and into drug development and clinical practice ^54,157^, and we dedicate the following sections to a reflection on these scientific, technical, and regulatory challenges and solutions which we believe are critical in developing clinically-relevant biomarkers of SUDs.

### Regulatory validation of neuroimaging biomarkers

The use of neuroimaging biomarkers in clinical and drug development contexts is contingent on approval by relevant regulatory bodies. These include the FDA in the US and the EMA in the European Union, which in recent years have developed structured frameworks within which biomarkers can be approved and endorsed for use, primarily in drug development and clinical trials ^158,159^. In the US, the 21st Century Cures Act adopted the process of qualification of drug development tools (including biomarkers) into US law in December 2016. Before the establishment of the drug development tool qualification program, FDA acceptance of biomarkers as drug development tools happened on a sponsor-by-sponsor, drug-by-drug basis. Biomarkers qualified under the current framework can be used by drug developers for the qualified context of use. Neuroimaging biomarkers submitted for approval through the FDA framework (and with some differences, the EMA framework) should be precisely defined with descriptions of the neuroimaging protocol, target populations, and the use context for which the biomarker is to be approved.

During the validation process, a biomarker’s analytical characteristics, such as reliability, validity, and natural variation need to be established. This is particularly important since despite some supporting evidence ^160^, there are significant concerns about the reliability of commonly used neuroimaging paradigms ^161^. Such research could also aid in the choice of biomarker: For example, a recent fMRI alcohol cue-reactivity study demonstrated that brain activations during constituting contrast conditions ‘alcohol’ and ‘neutral’ have higher reliability than the ‘alcohol versus neutral’ difference contrast ^143^. After analytical validation, the biomarker should be “clinically validated” by elucidating its etiological link to an SUD and establishing that it is reliably associated with current or future disease or recovery, for example by presenting evidence of the existence and role of neural aberrations in SUDs as was attempted in this manuscript. Finally, it should be demonstrated that the biomarker addresses a substantial gap and demonstrates cost-effectiveness. As an example of how these requirements can be met for a putative neuroimaging marker, Textbox 1 includes a brief discussion of the relevant evidence and important gaps in the case of markers of striatal structure and function. Besides these formal qualification pathways, the use of biomarkers in clinical contexts can be facilitated by the endorsement of a constellation of other institutions which develop relevant guidelines and best practice recommendations for SUDs. Meeting qualification standards for neuroimaging biomarkers requires broad collaboration and public-private partnerships, extensive resource sharing, and rigorous research practices. These qualification steps are outlined in Figure 7.

**Figure 7:**
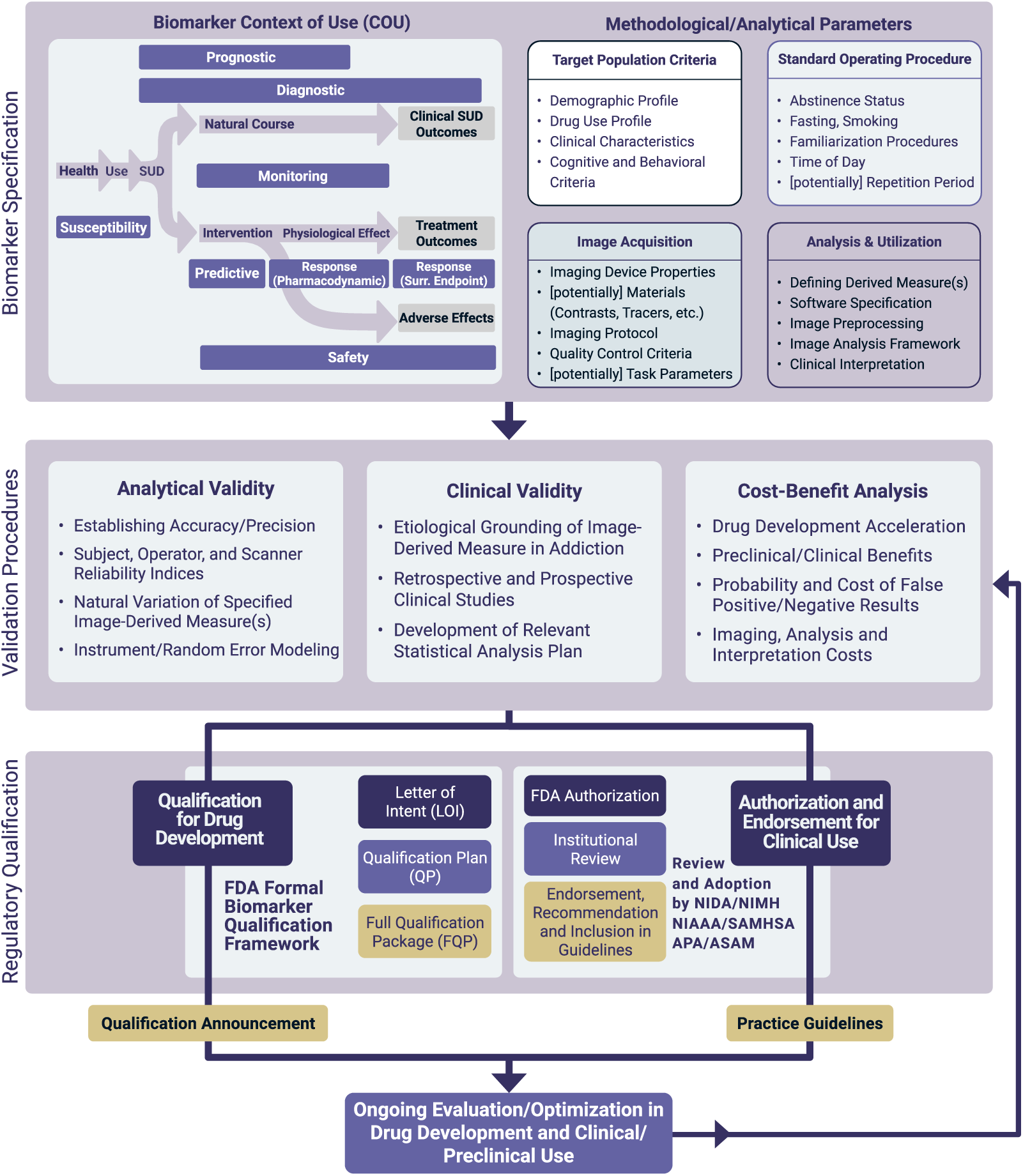
Major steps in the development and validation of potential neuroimaging biomarkers for SUDs. Initially, the context(s) of use for the biomarker is specified and the potential biomarker is precisely defined. Following analytical and clinical validation and cost-benefit analysis, the compiled evidence is presented for regulatory approval. The FDA evaluates the use of biomarkers for drug development through a biomarker qualification process involving submission of a Letter of Intent, a Qualification Plan, and a Full Qualification Package, though a Letter of Support may be issued by the FDA to indicate its support for a biomarker before formal qualification. The use of neuroimgaing biomarkers in clinical contexts also requires initial approval by the FDA, but also the endorsement of a constellation of other institutions (adapted from ^157^, reproduced with permission). Surr. Endpoint: Surrogate Endpoint, COU: Context of Use.

### Large-scale collaboration and multiple stakeholders in biomarker development

The development, validation, and impactful use of neuroimaging biomarkers of SUDs will depend upon the formation of large, multi-site consortia which can effectively direct resources towards biomarker discovery with harmonized research designs, starting with the “low-hanging fruit”-biomarkers with substantial bodies of supporting evidence and greatest potential utility, such as in intervention development. Furthermore, while translational research in the field is mostly conducted by academics, the developed biomarkers need to be cost-effective from the perspective of policymakers interested in reducing the societal burden of SUDs, pharmaceutical companies developing interventions and seeking to reduce the duration and cost of drug development, and regulatory bodies interested in using scientifically validated neuroimaging markers in approval decisions ^162^.

Importantly, the use of neuroimaging biomarkers and the interventions that they are used to develop should also be acceptable, accessible, affordable and desirable for individuals with SUDs, and concerns about neuroscientific models of addiction should be addressed^22^. Multi-stakeholder engagement is complicated by gaps in knowledge and terminology between stakeholders, differences in expectations and interests, power imbalances and stigma associated with SUDs, and identifying representative stakeholders. Effective engagement of various stakeholders in biomarker development for SUDs requires designing engagement plans and collaboration roadmaps, developing common terminology, clarifying and communicating the purpose of the engagement and stakeholder roles, and investing in the necessary skills and resources ^163–165^.

### Rigorous research and reporting for biomarker discovery

An essential step in the development of neuroimaging biomarkers is to harmonize best practices in study design, analysis, and reporting, especially given recent concerns about the reliability of multiple neuroimaging modalities ^166–168^. While there is significant disagreement over the best neuroimaging research design practices, certain factors would likely improve overall methodological quality ^18^. Larger sample sizes and appropriate statistical power analyses, for example, would improve the reproducibility of fMRI cue reactivity studies and enable the ascertainment of substantive effects ^169^. One solution is the collation of neuroimaging data into “big data” repositories, such as the structural MRI database maintained by the NeuroImaging Genetics through Meta-Analyses (ENIGMA) International Consortium ^170^ and task-based fMRI datasets made available on platforms such as OpenNeuro ^171^, which can be used for large-scale analyses, hypothesis generation, and model validation. A growing number of multi-center initiatives such as the Human Connectome Project, UK Biobank, and the Adolescent Brain Cognitive Development (ABCD) project collect neuroimaging data from thousands of individuals using harmonized scanning and data management standards across sites and may provide highly useful for the identification of neuroimaging markers ^172–174^. In the absence of large-scale studies, meta-analyses can be used to synthesise data across neuroimaging studies, discover convergent findings that replicate across SUDs and contexts of use, and disambiguate the influence of study design and confounders. A summary of neural markers which have replicated across SUDs or contexts of use is presented in Table 1.

Another issue is the methodological heterogeneity of neuroimaging research. The choice of hardware, data acquisition protocol, pre-processing steps, and analysis pipelines can have unexpected and substantial effects on the results of studies using a variety of neuroimaging modalities ^175–177^. While it is impossible to prescribe a similar set of best practices for every study, the design should be appropriate to specific contexts of use if the results are to contribute to biomarker development. Furthermore, the clarity, interpretability, and replicability of neuroimaging research would be enhanced with pre-registered protocols, carefully considering essential aspects of research design, and comprehensive reporting of methodological details ^178^. Various guidelines for research design and reporting have been developed in recent years with various degrees of generality, such as those developed by the Committee on Best Practice in Data Analysis and Sharing (COBIDAS) and COBIDAS MEEG ^179,180^ and the Addiction Cue Reactivity Initiative (ACRI) of the addiction working group of ENIGMA consortium ^54^.

### Technological advancements relevant to SUD biomarker discovery

A variety of innovations in neuroimaging technology, data management, and analysis may pave the way for SUD neuroimaging biomarkers. Among promising advances are high-field MRI with increasingly stronger magnetic fields, which can offer greater spatial resolution in structural and functional scans ^181^; functional magnetic resonance spectroscopy (fMRS), which can capture dynamic changes in metabolites ^182^; and new PET radiotracers, which can probe under-investigated neurotransmitter systems of interest to addiction medicine ^183^. Another emerging possibility is the use of neuroimaging to derive subject-specific “fingerprints” of brain circuitry or function, such as “precision functional mapping” to identify individual-level functional connectomes with fMRI ^184^ or the use of EEG to identify participant-specific electrophysiological patterns ^185,186^. Such subject-level (rather than group-level) neuroimaging markers are particularly useful for biomarker development since most contexts of use require biomarkers which can be used to make decisions for individual patients, and the heterogeneity of brain structure and function across individuals renders the translation of group-level findings to the individual-level problematic ^187,188^.

It is also increasingly possible to integrate different neuroimaging technologies concurrently or in series, and use multimodal data to probe multiple facets of brain structure and function in tandem: resting-state fMRI and MRS can be utilized together to assess the relationship between neuromodulation-associated brain network changes and neurotransmitter concentrations ^189^, functional diffuse correlation spectroscopy and functional near-infrared spectroscopy have been used along with EEG during and after brain stimulation to concurrently measure cerebral hemodynamics and electrical activity (Giovannella et al., 2018), simultaneous EEG and fMRI neurofeedback might improve the quality of the provided neurofeedback using bimodal data (Lioi et al., 2020), and receptor maps obtained by PET can inform resting-state fMRI functional connectivity analysis ^190^. These technological advances have co-occurred with rapid developments in informatics, data analytics, and computational infrastructure which facilitate data storage and sharing, biomarker discovery with increasingly sophisticated machine learning algorithms, and reproducible analytical practices ^191,192^.

### Theories and models of addiction

A significant challenge in biomarker development and theoretical progress in both addiction medicine and psychiatry as a whole is the fact that the DSM is a descriptive diagnostic manual, and its constructs are neither domain-based nor necessarily grounded in neurobiology ^193^. SUDs are multifaceted disorders with complex comorbidity patterns and overlapping brain substrates ^194,195^, and neuroimaging biomarkers will likely reflect the trans-diagnostic impairment and recovery of physiological processes which undergird specific cognitive domains. This highlights the importance of mechanistic models of disease (rather than manual-based diagnostic labels) in the development of neuroimaging addiction biomarkers. Under most mechanistic accounts of addiction, addiction starts with positive reinforcement learning before other processes are involved ^7^. These include excessive incentive sensitization ^139^ for example, which can explain heightened reactivity to drug cues in functional neuroimaging studies ^96,196–200^. What happens later is subject to some contention: some emphasize a shift from initially goal-directed behavior to habitual and then compulsive substance use, reflected in neuroimaging findings of a shift in drug cue-reactivity from the ventral to the dorsal striatum ^201^; while others highlight a shift from positive to negative reinforcement as withdrawal becomes more important, with some emphasizing goal-directed choice (rather than habit or compulsion) as individuals learn to relieve negative affect with substance use.

Other models focus on processes such as learning and executive control. The reward deficiency and allostasis models ^202^, for example, highlight the importance of suppression and disruption of reward processing circuits; while others focus on core deficits in value updates and reward learning ^203^. These models can explain wide-spread neural aberrations when individuals with various SUDs process non-drug gains and losses ^136^ and the reduced salience of novel and surprising stimuli ^204^. While the frameworks discussed above can account for frequent observations of impaired response inhibition^205^ (and corresponding neuroimaging aberrations during executive control tasks ^198,206–209^), recent “dual process” accounts of addiction emphasize the broad disruption of top-down, deliberative processes in prefrontal and parietal regions together with deregulation and disinhibition of bottom-up automatic processes in mesolimbic circuits ^210^. Further, recent observations suggest that general cognitive decline and a broad depletion of executive control in addiction may be particularly important to the course of disease and treatment ^211^, in line with broad degenerations of cortical gray and white matter ^39,41,43,47,212^. It must be emphasized that many of these contructs are not mutually exclusive, and multiple interacting processes may be in play in the development, maintenance and recovery from SUDs.

Overall, the briefly discussed models (see ^10^ for detailed discussion) and theories have been developed in tandem with advances in addiction neuroimaging, and provide promising starting points for the development of neuroimaging biomarkers. Frameworks such as the impaired response inhibition and salience attribution model ^11^, the Addictions Neuroclinical Assessment framework ^8^, and the Alcohol and Addiction Research Domain Criteria ^213^ aim to map addictive disorders to specific axes of impairment and neuroimaging research by facilitating hypothesis generation and the development of interpretable neuroimaging biomarkers linked to formal theories of addiction. Despite differences, these frameworks converge on the involvement of positive valence, negative valence and cognitive control systems in SUDs, and have been used to propose neuroscience-informed classifications of interventions ^10,214^. Complementing these theoretical developments, computational modeling of processes of interest in addiction neuroscience (such as drug cue-reactivity, aberrant decision-making, etc.) can mechanistically represent the interplay between neural mechanisms and behavior and link neuroimaging markers, underlying neuro-cognitive pathology, and signs and symptoms of SUDs ^215,216^.

## CONCLUSION

Modern neuroimaging technologies can probe brain structure and function at unprecedented resolution and have already produced novel insights into the neurocognitive mechanisms of addiction and recovery. The rapid pace of technological advancement, increasing availability, and growing recognition of neuroimaging paradigms in recent years has contributed to an explosion in their use within clinical and translational addiction medicine: from 2015 to 2021, an average of 35 protocols with neuroimaging as one of the registered outcome measures in people with SUDs were registered on ClinicalTrials.gov every year, more than ten times the average number from 2000 to 2006. Especially popular are fMRI (268 protocols) and EEG (50 protocols), which dynamically assess brain function; PET (71 protocols) and MRS (35 protocols) which probe neurotransmitter systems and their interactions with radioligands; and structural MRI (35 protocols) which can be used to investigate brain structure at various scales. These paradigms can be systematically utilized to discover and develop biomarkers, measures that objectively reflect biological processes involved in both the progression of substance use and SUDs and the physiological and clinical impact of interventions for these disorders. Particularly promising are several neuroimaging markers which have replicated in meta-analyses across contexts of use and disorders. Technological and scientific advancements, rigorous research practices, and multi-stakeholder engagement can facilitate the development of institutionally approved neuroimaging biomarkers that enable impactful, personalized interventions for SUDs to be used in clinical practice in the foreseeable future.

## METHODS AND LIMITATIONS

The present manuscript is informed by two systematic reviews. The first covered SUD clinical research protocols which include neuroimaging outcome measures, obtained by querying the ClinicalTrials.gov repository between inception and November 17, 2021 (Supplementary Figure 1a). This systematic review yielded a final result of 409 protocols. The second systematic review was conducted on PubMed, focusing on meta-analyses of neuroimaging studies of SUDs and finding 61 meta-analyses from which 83 meta-analytic findings were extracted (Supplementary Figure 1b). Please refer to the methods section of the supplementary materials for more details on the methods, and to the OSF repository https://osf.io/79uc3/?view_only=1d92a6fd769f40119464b156f0c88912 for the search protocol and analysis scripts. Although we used widely-known and inclusive databases of protocols and meta-analyses, we did not triangulate the results with other databases. Our approach likely leads to some missing protocols and papers, and in particular an under-representation of protocols from countries that use registration platforms other than ClinicalTrials.gov.

## Data Availability

The protocol and data for this systematic review are available on the open science framework (OSF) website (https://osf.io/79uc3/?view_only=1d92a6fd769f40119464b156f0c88912).
The ClinicalTrials.gov search engine was used through the Study Fields query URL (https://ClinicalTrials.gov/api/gui/ref/api_urls) for searching the clinical trial protocols.
For full-text screening, all available records were downloaded from the Aggregate Analysis of ClinicalTrials.gov (AACT) Database, Clinical Trials Transformation Initiative (CTTI) database 217 (https://aact.ctti-clinicaltrials.org/) for the second stage. For searching the systematic reviews and meta-analyses, studies were identified using the Medline/PubMed (https://pubmed.ncbi.nlm.nih.gov/) database.

https://osf.io/79uc3/?view_only=1d92a6fd769f40119464b156f0c88912

## DATA AVAILABILITY STATEMENT

The protocol and data for this systematic review are available on the open science framework (OSF) website (https://osf.io/79uc3/?view_only=1d92a6fd769f40119464b156f0c88912). The ClinicalTrials.gov search engine was used through the Study Fields query URL (https://ClinicalTrials.gov/api/gui/ref/api_urls) for searching the clinical trial protocols. For full-text screening, all available records were downloaded from the Aggregate Analysis of ClinicalTrials.gov (AACT) Database, Clinical Trials Transformation Initiative (CTTI) database ^217^ (https://aact.ctti-clinicaltrials.org/) for the second stage. For searching the systematic reviews and meta-analyses, studies were identified using the Medline/PubMed (https://pubmed.ncbi.nlm.nih.gov/) database.

## CODE AVAILABILITY STATEMENT

All codes are available on the study’s OSF project repository at the following link: https://osf.io/79uc3/?view_only=1d92a6fd769f40119464b156f0c88912 Data analyses and illustrations were conducted using R version 4.0.5 ^218^, with dplyr ^219^ and ggplot2 ^220^ packages. The codes for data illustrations are freely available on the OSF repository of this project.

## ACKNOWLEDGEMENT

H.E. is supported by funds from the Laureate Institute for Brain Research and Medical Discovery Team on Addiction and Brain Behavior Foundation (NARSAD Young Investigator Award 27305). O.C. is funded by NIH grants AG07425801, AG077497, AG077000, AG067765, AG041200, AG062309, AG062200, and AG069476, William K. Warren Foundation and the National Institute of General Medical Sciences Center Grant Award Number (1P20GM121312) and the National Institute on Drug Abuse (U01DA050989). A.R.C. is funded by NIH/NIDA mechanisms UG1 DA050209, R01DA039215,T32-DA-028874, P30 DA046345, and U01DA048517. T.R. was substantially involved in UG1DA050209 and U01DA048517 consistent with her role as Scientific Officer. She has no substantial involvement in the other cited grants.

## AUTHORS CONTRIBUTIONS STATEMENT

Kathleen Brady, Owen Carmichael, Anna Rose Childress, Hamed Ekhtiari, F. Gerard Moeller, Patricia O’Donnell, Maria Oquendo, Martin Paulus, Diego Pizzagalli, Tanya Ramey, and Joseph Schacht conceived of the presented idea and designed the study and Hamed Ekhtiari coordinated the consensus process among all authors. Arshiya Sangchooli, Mehran Zare-Bidoky, and Hamed Ekhtiari gathered data, designed the tables, and performed the initial analytic calculations. All authors discussed the results and contributed to the final manuscript.

## COMPETING INTERESTS STATEMENT

O.C. has received grant funding from Eli Lilly, Inc, and Nestle, Inc. He has provided paid consulting to Novo Nordisk. Dr. Paulus is an advisor to Spring Care, Inc., a behavioral health startup, he has received royalties for an article about methamphetamine in UpToDate. M.P.P. has a consulting agreement with and receives compensation from F. Hoffmann-La Roche Ltd. P.O. is an employee and shareholder of Sage Therapeutics. Other authors report no conflicts of interest.

Disclaimer: The views and opinions expressed in this manuscript are those of the authors only and do not necessarily represent the views, official policy or position of the U.S. Department of Health and Human Services or any of its affiliated institutions or agencies.

