## Supplementary Materials for "Neuroimaging Biomarkers in Addiction"

**Note:** To reiterate and avoid confusion, our 409 protocols contained 479 different neuroimaging modalities, such as structural MRI, functional MRI, and EEG. With a more fine-grained classification of neuroimaging paradigms into subclasses (e.g., splitting fMRI into resting-state and task types), the 409 protocols had 688 neuroimaging outcome measures. On the other hand, a few protocols did not use any neuroimaging modalities in a way that fit any biomarker context of use while some protocols employed neuroimaging modalities within multiple contexts of use, leading to 520 distinct uses of neuroimaging modalities in different biomarker categories across our 409 protocols.

### METHODS

#### Systematic Review of Protocols on ClinicalTrials.gov

The methods section is organized based on the Preferred Reporting Items for Systematic Reviews and Meta-Analyses (PRISMA) checklist [5].

**Eligibility criteria:** Any study or trial protocol registered on the ClinicalTrials.gov repository was included based on the criteria listed below:

**Study design and methodology:** Of interest were studies that include one or more neuroimaging modalities as a measure in their methods, whether for observational purposes, as an intervention outcome measure, or as a treatment tool. We mainly focused on SUDs and not the other types of addiction like video gaming and gambling; therefore, types of addiction other than SUDs are excluded from this study. The neuroimaging investigation had to be conducted for at least some of the study population. We did not exclude protocols of the studies based on status; i.e., even terminated, withdrawn, or suspended protocols were included to minimize reporting bias.

**Participants:** Every study required at least one human population or sub-population with more than one individual, for at least some of whom one of the following needed to be true:

1. At least one circumscribed group of participants had a diagnosis of at least one substance use disorder which is active, relapsed, or in remission; with the diagnosis made either before the study, as part of the study protocol during the investigation, or by the end of the study (i.e., with the diagnosis serving as an outcome measure).

2. At least one group of participants was included explicitly because they regularly consume a substance and/or had a risky pattern of consumption that might lead to addiction and the study assessed them with neuroimaging modalities.

3. The subjects were not diagnosed with SUDs but the study aimed to investigate the addiction/dependence-related impacts of a substance that is of interest in addiction medicine (e.g., alcohol, methamphetamine) using neuroimaging.

For instance, the following protocol was excluded since only the pain-relieving effects of morphine were investigated, rather than its addictive potential: https://clinicaltrials.gov/show/NCT01878006. Interventional studies which used neuroimaging on people with SUDs to assess the effects of an intervention on aspects unrelated to SUDs were excluded. For example, the following protocol was excluded since its focus is the effect of an intervention on depression among people with SUDs which is arguably not directly related to treating SUDs: https://clinicaltrials.gov/show/NCT02189915. No restriction was placed on study participants based on demographic, biological, or clinical factors (such as any comorbidities).

**Information source:** Sources were identified and retrieved among protocols registered on the ClinicalTrials.gov repository. Relevant protocols were identified using a comprehensive search strategy for all terms related to SUDs and neuroimaging as outlined below. The Study Fields query URL type (https://ClinicalTrials.gov/api/gui/ref/api_urls) was used to query and download the information. All available columns of the data were downloaded into a comma-separated file for screening. For full-text screening, all available records were downloaded from the Aggregate Analysis of ClinicalTrials.gov (AACT) Database, Clinical Trials Transformation Initiative (CTTI) database [1] (https://aact.ctti-clinicaltrials.org/) for the second stage of screening, and fields that would not be relevant to the screening were removed.

**Search strategy**: Considering the subject of the review, a list of two sets of keywords was compiled by H.E., A.S., and M.Z.B. These terms were adapted for use in the ClinicalTrials.gov repository, and the exact search syntax is outlined in Supplementary Table 1. The first set includes types of neuroimaging modalities and the second includes terms related to substance use. To help widen the search, no filters were used.

**Data management**: Search results were imported to Microsoft Excel (Professional Plus 2019) [6]. Screening of articles for relevance was performed by reviewing the full webpage of the study protocols. All raw and processed data tables were version-tracked, with the latest version available on the OSF repository.

**Study selection:** Screening forms were developed and studies were checked by two authors (A.S. and M.Z.B.). The authors initially checked the eligibility of fifty randomly chosen studies under the supervision of the corresponding author (H.E.), as a calibration exercise to ensure eligibility criteria are applied consistently [7]. Any protocols with an uncertain eligibility status after full-text screening were then discussed with H.E. until a consensus on their inclusion was reached. Reasons for the exclusion of study protocols were recorded, according to the PRISMA framework [8]. Neither of the review authors was blind to the protocol authors or institutions.

**Data collection:** Data were filled into an electronic spreadsheet by A.S and M.Z.B. Consistency between the authors was honed through a calibration exercise in which all authors evaluated and discussed their ratings for 20 randomly chosen studies [7]. A.S., M.Z.B., and H.E. further refined the data extraction form to reduce inconsistency and ambiguity after the exercise. Each article was reviewed independently by the two authors in two separate spreadsheets, with inconsistencies resolved in discussions with H.E.

**Data items:** We extracted protocol details, including the substance (main substance(s) of interest in the study), main protocol type (whether the protocol was for an observational or interventional study), imaging modality (e.g., MRI, PET), functional paradigm (for functional modalities such as fMRI or EEG, whether the imaging was during resting-state or task performance), task category if applicable, category and subcategory of interventions included in each protocol, study start date, and basic participant characteristics. For each protocol, we also extracted the context(s) of use in which each utilized neuroimaging modality could contribute to biomarker development, for one or more substance use disorders. If a study did not contain enough information to extract or infer putative biomarker contexts of use, biomarker context of use was rated "not-specified".

The full list of the final studies and their characteristics can be found on the OSF repository.

**Software:** The ClinicalTrials.gov search engine was used through the Study Fields query URL (https://ClinicalTrials.gov/api/gui/ref/api_urls). The AACT CTTI database (https://aact.ctti-clinicaltrials.org/) was used to extract structured protocol data for the second screening stage. Google Sheets from Google's Google Docs Editors suite was used to design tables for data extraction and share them among authors. Microsoft Excel was used for data storage and ratings. Data analyses and illustrations were conducted using R version 4.0.5 [2], with dplyr [3] and ggplot2 [4] packages. The codes for data illustrations are freely available on the OSF repository of this project.

The protocol for this systematic review was developed throughout 2021, and was first uploaded on GitHub and uploaded to the OSF repository in 2022. The latest version of the systematic review protocol and the database of included studies can be viewed in the repository.

**Risk of Bias Assessment:** Since this review focused on all the protocols registered on ClinicalTrials.gov at the beginning of each study, this review is less prone to the risk of bias in reporting.

**Synthesis:** Given the breadth of the review, the heterogeneity of the protocols, and that most protocols do not contain quantitative results, no quantitative synthesis was planned for this systematic review. Cross-tabulations and a Sankey diagram are used to present the relationship between the categorical variables in the study, and the synthesis is otherwise narrative.

***Supplementary Table 1:*** *Terms used to systematically search for neuroimaging studies in addiction.*

| | **Search Terms for ClinicalTrials.gov** | **Search Terms for MedLine/PubMed** |
|---|---|---|
| **Addiction** | AREA[ConditionSearch] (addicted OR addiction OR dependence OR “substance use” OR “substance abuse” OR “drug misuse” OR “substance-related disorder” OR intoxication OR “drug withdrawal” OR “drug abuse” OR “drug use” OR smoker OR cigarette OR cocaine OR methamphetamine OR marijuana OR cannabis OR alcohol OR alcoholism OR heroin) | addicted OR addiction OR dependence OR "substance use" OR "substance abuse" OR "drug misuse" OR "substance-related disorder" OR intoxication OR "drug withdrawal" OR "drug abuse" OR "drug use" OR smoker OR cigarette OR cocaine OR methamphetamine OR marijuana OR cannabis OR alcohol OR alcoholism OR heroin |
| **Neuroimaging** | AREA[OutcomeSearch] (MRI OR PET OR tracer OR glutamate OR MRS OR spectroscopy OR EEG OR electroencephalography OR EcoG OR SPECT OR MEG OR acetylaspartate OR DSC OR DCE OR ASL OR fNIRS OR “functional ultrasound” OR “arterial spin” OR fUS OR “blood oxygen level” OR fMRI OR hemodynamic OR BOLD OR “event related” OR diffiusion OR DTI OR signal OR evoked OR ERP OR accumbens OR insula OR parietal OR synchrony OR synchronous OR neuroinflammation OR neurovascular OR structural OR volumetry OR cingulate OR frontal OR prefrontal OR amygdala OR DMN OR ECN OR limbic OR task OR resting OR brain OR neural OR cortex OR connectivity OR circuitry OR circuit OR receptor OR occupancy OR binding OR imaging OR activation OR neuro) | MRI OR PET OR tracer OR glutamate OR MRS OR spectroscopy OR EEG OR electroencephalography OR ECoG OR SPECT OR MEG OR acetylaspartate OR DSC OR DCE OR ASL OR fNIRS OR "functional ultrasound" OR "arterial spin" OR fUS OR "blood oxygen level" OR fMRI OR hemodynamic OR BOLD OR "event related" OR diffusion OR DTI OR signal OR evoked OR ERP OR accumbens OR insula OR parietal OR synchrony OR synchronous OR neuroinflammation OR neurovascular OR structural OR volumetry OR cingulate OR frontal OR prefrontal OR amygdala OR DMN OR ECN OR limbic OR task OR resting OR brain OR neural OR cortex OR connectivity OR circuitry OR circuit OR receptor OR occupancy OR binding OR imaging OR activation OR neuro |
| **Full search** | Addiction-related AND Neuroimaging-related terms | Addiction-related AND Neuroimaging-related terms |

### Systematic Review of Meta-Analyses

The methods section is organized based on the Preferred Reporting Items for Systematic Reviews and Meta-Analyses (PRISMA) checklist. In this study, we only focused on substance use disorder (SUDs) and excluded other potential types of addiction like gaming or gambling.

**Eligibility criteria:** Meta-analyses studies were selected according to the criteria outlined below.

**Study design and methodology:** Of interest were Peer-reviewed meta-/mega-analyses that included one or more neuroimaging investigations as a major part of their assessment which at least one of the assessments directly fall into one of the FDA biomarker categories (refer to the introduction section for their definitions)

**Participants**: Every study required at least one human population or sub-population with more than one member with the use or diagnosis of SUD at the beginning, during or at the end of the study.

**Information Source:** Studies were identified using the Medline/PubMed (https://pubmed.ncbi.nlm.nih.gov/) database. Relevant articles were identified using the same search strategy used for protocol search combined with relevant search terms for meta-analysis and systematic reviews (Supplementary Table 1). The search results were downloaded in comma-separated format and the required data including author information, doi, PMID, article title, and publication year were entered into an Excel spreadsheet.

**Language:** Only publications with their full text in English were included.

**Data management:** Search results were imported to Excel. Screening of articles for relevance was performed by first reviewing the Title and Abstract and then the full text of the eligible articles. All raw and processed data tables were version-tracked, with the latest version available on the OSF repository.

**Study selection:** Screening forms were developed and studies were checked by two authors (A.S. and M.Z.B.). The authors initially checked the eligibility of fifty randomly chosen studies under the supervision of the corresponding author (H.E.), as a calibration exercise to ensure eligibility criteria are applied consistently [7]. Any protocols with an uncertain eligibility status after full-text screening were then discussed with H.E. until a consensus on their inclusion was reached. Reasons for the exclusion of study protocols were recorded, according to the PRISMA framework (Moher et al., 2015). Neither of the review authors was blind to the protocol authors or institutions.

**Data collection:** Data were filled into an electronic spreadsheet by A.S. and M.Z.B. Consistency between the authors was honed through a calibration exercise in which all authors evaluated and discussed their ratings for 10 randomly chosen studies [7]. A.S., M.Z.B., and H.E. further refined the data extraction form to reduce inconsistency and ambiguity after the exercise. Each article was reviewed independently by the two authors in two separate spreadsheets, with inconsistencies resolved in discussions with H.E.

**Data items:** We extracted meta-analysis details, including the number of studies and participants in the entire meta-analysis, the number of studies and participants in each analysis, the substance (main substance(s) of interest in the study), imaging modality (e.g., MRI, PET), functional paradigm (for functional modalities such as fMRI or EEG, whether the imaging was during resting-state or task performance), task category if applicable. For each study, we also extracted the context(s) of use in which each utilized neuroimaging modality could contribute to biomarker development, for one or more SUDs and the putative biomarker role neuroimaging had in that analysis.

***Supplementary Table 2:*** *References for the putative neuroimaging biomarkers in Figure 7. For each marker, a paper demonstrating aberrations in SUDs is presented alongside a clinical from the systematic review database using this marker.*

| **Neuroimaging Marker** | **Substance** | **Clinical Trial** | **Registration Number** | **Related Study** |
|---|---|---|---|---|
| Late positive potential | Cocaine | [9] | NCT04994821 | [10] |
| Prefrontal-striatal FC | Methamphetamine | [11] | NCT03748875 | [12] |
| DLPFC glutamate | Alcohol | [13] | NCT03291431 | [14] |
| Acetylcholine receptors | Nicotine | [15] | NCT01526005 | [16] |
| Hippocampal volume | Cannabis | [17] | NCT04902092 | [18] |
| Striatal cue reactivity | Alcohol | [19] | NCT04366505 | [20] |
| Corpus callosal integrity | Cocaine | [21] | NCT02774343 | [22] |

***Supplementary Table 3:*** *Results of the systematic review of meta-analyses. Findings from the 61 meta-analyses are broken down into 83 rows, each corresponding to a marker with a specific context of use for a substance use disorder.* dMRI: diffusion magnetic resonance imaging ; EEG: electroencephalography; *fMRI: functional* magnetic resonance imaging; fNIRS: Functional near-infrared spectroscopy; MEG: magnetoencephalography; *MRI:* magnetic resonance imaging; *MRS: magnetic resonance-spectroscopy; PET:* positron emission tomography; *sMRI: structural* magnetic resonance imaging; *SPECT: Single-photon emission computed tomography;*

| Title | Year | Substance Use Disorder(s) | Biomarker Type | Scale | Modality | Task |
|---|---|---|---|---|---|---|
| Association of Stimulant Use With Dopaminergic Alterations in Users of Cocaine, Amphetamine, or Methamphetamine: A Systematic Review and Meta-analysis | 2017 | Methamphetamine/ Cocaine | Diagnostic | Biochemical | PET/ SPECT | NA |
| Association of Stimulant Use With Dopaminergic Alterations in Users of Cocaine, Amphetamine, or Methamphetamine: A Systematic Review and Meta-analysis | 2017 | Methamphetamine/ Cocaine | Diagnostic | Biochemical | PET/ SPECT | NA |
| Association of Stimulant Use With Dopaminergic Alterations in Users of Cocaine, Amphetamine, or Methamphetamine: A Systematic Review and Meta-analysis | 2017 | Methamphetamine/ Cocaine | Diagnostic | Biochemical | PET/ SPECT | NA |
| Regional differences in white matter integrity in stimulant use disorders: A meta-analysis of diffusion tensor imaging studies | 2019 | Stimulants | Diagnostic | Structural | dMRI | NA |
| Regular cannabis use is associated with altered activation of central executive and default mode networks even after prolonged abstinence in adolescent users: Results from a complementary meta-analysis | 2019 | Cannabis | Diagnostic | Hemodynamic | fMRI | Multiple |
| Regular cannabis use is associated with altered activation of central executive and default mode networks even after prolonged abstinence in adolescent users: Results from a complementary meta-analysis | 2019 | Cannabis | Response | Hemodynamic | fMRI | Multiple |

| | | | | | | |
|---|---|---|---|---|---|---|
| Residual effects of cannabis use in adolescent and adult brains – a meta-analysis of fMRI studies | 2018 | Cannabis | Diagnostic | Hemodynamic | fMRI | Multiple |
| Residual effects of cannabis use in adolescent and adult brains – a meta-analysis of fMRI studies | 2018 | Cannabis | Diagnostic | Hemodynamic | fMRI | Multiple |
| The brain activity pattern in alcohol-use disorders under inhibition response Task | 2023 | Alcohol | Diagnostic | Hemodynamic | fMRI | Response Inhibition |
| How the harm of drugs and their availability affect brain reactions to drug cues: a meta-analysis of 64 neuroimaging activation studies | 2020 | General | Diagnostic | Hemodynamic | fMRI/ PET | Cue-Reactivity |
| How the harm of drugs and their availability affect brain reactions to drug cues: a meta-analysis of 64 neuroimaging activation studies | 2020 | General | Response | Hemodynamic | fMRI/ PET | Cue-Reactivity |
| Nature of glutamate alterations in substance dependence: A systematic review and meta-analysis of proton magnetic resonance spectroscopy studies | 2021 | General | Diagnostic | Biochemical | MRS | NA |
| Disrupted functional connectivity of the brain reward system in substance use problems: A meta-analysis of functional neuroimaging studies | 2023 | General | Diagnostic | Hemodynamic | fMRI | Rest |
| Neural substrates of smoking cue reactivity: a meta-analysis of fMRI studies | 2012 | Nicotine | Response | Hemodynamic | fMRI | Cue-Reactivity |
| The P300 event-related brain potential as a neurobiological endophenotype for substance use disorders: a meta-analytic investigation | 2012 | General | Diagnostic | Electrophysiology | EEG | Rest |
| Alcohol and Neural Dynamics: A Meta-analysis of Acute Alcohol Effects on Event-Related Brain Potentials | 2021 | Alcohol | Response | Electrophysiology | EEG | Multiple |
| Regional cerebral blood flow predictors of relapse and resilience in substance use recovery: A coordinate-based meta-analysis of human neuroimaging studies | 2018 | General | Predictive | Hemodynamic | fMRI/ PET | Multiple |
| Predicting alcohol dependence from multi-site brain structural measures | 2022 | Alcohol | Diagnostic | Structural | sMRI | NA |

| | | | | | | |
|---|---|---|---|---|---|---|
| Gray matter abnormalities in cocaine versus methamphetamine-dependent patients: a neuroimaging meta-analysis | 2015 | Cocaine | Diagnostic | Structural | sMRI | NA |
| The P300 in alcohol use disorder: A meta-analysis and meta-regression | 2019 | Alcohol | Diagnostic | Electrophysiology | EEG | Multiple |
| Visual cortex activation to drug cues: a meta-analysis of functional neuroimaging papers in addiction and substance abuse literature | 2014 | General | Diagnostic | Hemodynamic | fMRI | Cue-Reactivity |
| Convergent gray matter alterations across drugs of abuse and network-level implications: A meta-analysis of structural MRI studies | 2022 | General | Diagnostic | Structural | sMRI | NA |
| Brain metabolite alterations related to alcohol use: a meta-analysis of proton magnetic resonance spectroscopy studies | 2022 | Alcohol | Diagnostic | Biochemical | MRS | NA |
| Common and separable neural alterations in substance use disorders: A coordinate-based meta-analyses of functional neuroimaging studies in humans | 2020 | Nicotine/ Alcohol/ Cocaine/ Cannabis | Diagnostic | Hemodynamic | fMRI | Multiple |
| Common and separable neural alterations in substance use disorders: A coordinate-based meta-analyses of functional neuroimaging studies in humans | 2020 | Nicotine | Diagnostic | Hemodynamic | fMRI | Multiple |
| Common and separable neural alterations in substance use disorders: A coordinate-based meta-analyses of functional neuroimaging studies in humans | 2020 | Alcohol | Diagnostic | Hemodynamic | fMRI | Multiple |
| Common neurofunctional dysregulations characterize obsessive-compulsive, substance use, and gaming disorders-An activation likelihood meta-analysis of functional imaging studies | 2021 | General | Diagnostic | Hemodynamic | fMRI | Multiple |
| Common abnormality of gray matter integrity in substance use disorder and obsessive-compulsive disorder: A comparative voxel-based meta-analysis | 2021 | General | Diagnostic | Structural | sMRI | NA |
| Common biology of craving across legal and illegal drugs - a quantitative meta-analysis of cue-reactivity brain response | 2011 | Nicotine/ Alcohol/ Cocaine | Diagnostic | Hemodynamic | fMRI/ PET | Cue-Reactivity |

| | | | | | | |
|---|---|---|---|---|---|---|
| Distinct patterns of prefrontal cortical disengagement during inhibitory control in addiction: A meta-analysis based on population characteristics | 2021 | General | Diagnostic | Hemodynamic | fMRI | Response Inhibition |
| Lower regional grey matter in alcohol use disorders: evidence from a voxel-based meta-analysis | 2021 | Alcohol | Diagnostic | Structural | sMRI | NA |
| Lower regional grey matter in alcohol use disorders: evidence from a voxel-based meta-analysis | 2021 | Alcohol | Response | Structural | sMRI | NA |
| Electrophysiological indices of biased cognitive processing of substance-related cues: a meta-analysis | 2012 | General | Diagnostic | Electrophysiology | EEG | Attention |
| Addiction as a brain disease? A meta-regression comparison of error-related brain potentials between addiction and neurological diseases | 2023 | General | Diagnostic | Electrophysiology | EEG | Error |
| Distinct brain structural abnormalities in attention-deficit/ hyperactivity disorder and substance use disorders: A comparative meta-analysis | 2022 | General | Diagnostic | Structural | sMRI | NA |
| Disruption of Reward Processing in Addiction : An Image-Based Meta-analysis of Functional Magnetic Resonance Imaging Studies | 2017 | General | Diagnostic | Hemodynamic | fMRI | Reward Task |
| White matter volume in alcohol use disorders: a meta-analysis | 2013 | Alcohol | Diagnostic | Structural | sMRI | NA |
| White matter volume in alcohol use disorders: a meta-analysis | 2013 | Alcohol | Response | Structural | sMRI | NA |
| Largely overlapping neuronal substrates of reactivity to drug, gambling, food and sexual cues: A comprehensive meta-analysis | 2016 | General | Diagnostic | Hemodynamic | fMRI/ PET/ SPECT | Multiple |
| Chronic smoking and brain gray matter changes: evidence from meta-analysis of voxel-based morphometry studies | 2013 | Nicotine | Diagnostic | Structural | sMRI | NA |
| Gray and white matter morphology in substance use disorders: a neuroimaging systematic review and meta-analysis | 2021 | General | Diagnostic | Structural | sMRI | NA |

| | | | | | | |
|---|---|---|---|---|---|---|
| Gray and white matter morphology in substance use disorders: a neuroimaging systematic review and meta-analysis | 2021 | General | Diagnostic | Structural | sMRI | NA |
| Functional neuroanatomy of craving in heroin use disorder: voxel-based meta-analysis of functional magnetic resonance imaging (fMRI) drug cue reactivity studies | 2023 | Opioids | Diagnostic | Hemodynamic | fMRI | Cue-Reactivity |
| Effects of stimulant drug use on the dopaminergic system: A systematic review and meta-analysis of in vivo neuroimaging studies | 2019 | Nicotine | Diagnostic | Biochemical | PET/ SPECT | NA |
| Effects of stimulant drug use on the dopaminergic system: A systematic review and meta-analysis of in vivo neuroimaging studies | 2019 | Cocaine | Diagnostic | Biochemical | PET/ SPECT | NA |
| Effects of stimulant drug use on the dopaminergic system: A systematic review and meta-analysis of in vivo neuroimaging studies | 2019 | Amphetamine | Diagnostic | Biochemical | PET/ SPECT | NA |
| Effects of stimulant drug use on the dopaminergic system: A systematic review and meta-analysis of in vivo neuroimaging studies | 2019 | Methamphetamine | Diagnostic | Biochemical | PET/ SPECT | NA |
| A voxel-wise meta-analysis of task-based functional MRI studies on impaired gain and loss processing in adults with addiction | 2021 | General | Diagnostic | Hemodynamic | fMRI | Loss and Gain |
| Altered neural activities during response inhibition in adults with addiction: a voxel-wise meta-analysis | 2021 | General | Diagnostic | Hemodynamic | fMRI | Response Inhibition |
| Common and gender-specific associations with cocaine use on gray matter volume: Data from the ENIGMA addiction working group | 2020 | Cocaine | Diagnostic | Structural | sMRI | NA |
| Is cannabis neurotoxic for the healthy brain? A meta-analytical review of structural brain alterations in non-psychotic users | 2013 | Cannabis | Diagnostic | Structural | sMRI | NA |
| Functional neuroimaging studies of alcohol cue reactivity: a quantitative meta-analysis and systematic review | 2013 | Alcohol | Diagnostic | Hemodynamic | fMRI/ PET | Cue-Reactivity |
| Functional neuroimaging studies of alcohol cue reactivity: a quantitative meta-analysis and systematic review | 2013 | Alcohol | Response | Hemodynamic | fMRI/ PET | Cue-Reactivity |

| | | | | | | |
|---|---|---|---|---|---|---|
| Spectroscopic meta-analyses reveal novel metabolite profiles across methamphetamine and cocaine substance use disorder | 2023 | Methamphetamine/ Cocaine | Diagnostic | Biochemical | MRS | NA |
| A coordinate-based meta-analysis of white matter alterations in patients with alcohol | 2022 | Alcohol | Diagnostic | Structural | dMRI/ sMRI | NA |
| Meta-analysis of grey matter changes and their behavioral characterization in patients with alcohol use disorder | 2021 | Alcohol | Diagnostic | Structural | sMRI | NA |
| A meta-analysis of tract-based spatial statistics studies examining white matter integrity in cocaine use disorder | 2021 | Cocaine | Diagnostic | Structural | dMRI | NA |
| Chronic cigarette smoking is linked with structural alterations in brain regions showing acute nicotinic drug-induced functional modulations | 2016 | Nicotine | Response | Hemodynamic | fMRI | Multiple |
| Neurobiological impact of nicotinic acetylcholine receptor agonists: an activation likelihood estimation meta-analysis of pharmacologic neuroimaging studies | 2015 | Nicotine | Response | Hemodynamic | fMRI/ PET | Multiple |
| Chronic cigarette smoking is linked with structural alterations in brain regions showing acute nicotinic drug-induced functional modulations | 2016 | Nicotine | Diagnostic | Structural | sMRI | NA |
| Shared network-level functional alterations across substance use disorders: A multi-level kernel density meta-analysis of resting-state functional connectivity studies | 2022 | General | Diagnostic | Hemodynamic | fMRI | Rest |
| Meta-analysis and review of functional neuroimaging differences underlying adolescent vulnerability to substance use | 2020 | General | Susceptibility | Hemodynamic | fMRI | Multiple |
| Neural representation of prediction error signals in substance users | 2021 | General | Diagnostic | Hemodynamic | fMRI | Multiple |
| Brain network dysfunctions in addiction: a meta-analysis of resting-state functional connectivity | 2022 | General | Diagnostic | Hemodynamic | fMRI | Rest |
| Late positive potential as a candidate biomarker of motivational relevance in substance use: Evidence from a meta-analysis | 2022 | General | Diagnostic | Electrophysiology | EEG | Cue-Reactivity |

| | | | | | | |
|---|---|---|---|---|---|---|
| Gray matter abnormalities in opioid-dependent patients: A neuroimaging meta-analysis | 2017 | Opioids | Diagnostic | Structural | sMRI | NA |
| Gray matter abnormalities in opioid-dependent patients: A neuroimaging meta-analysis | 2017 | Opioids | Response | Structural | sMRI | NA |
| White matter abnormalities in long-term heroin users: a preliminary neuroimaging meta-analysis | 2015 | Heroin | Diagnostic | Structural | dMRI | NA |
| White matter abnormalities in long-term heroin users: a preliminary neuroimaging meta-analysis | 2015 | Heroin | Response | Structural | dMRI | NA |
| Regional gray matter deficits in alcohol dependence: A meta-analysis of voxel-based morphometry studies | 2015 | Alcohol | Diagnostic | Structural | sMRI | NA |
| Functional and structural brain abnormalities in substance use disorder: A multimodal meta-analysis of neuroimaging studies | 2023 | General | Diagnostic | Structural | sMRI | NA |
| Functional and structural brain abnormalities in substance use disorder: A multimodal meta-analysis of neuroimaging studies | 2023 | General | Diagnostic | Hemodynamic | fMRI | Rest |
| Neuroimaging meta-analysis of cannabis use studies reveals convergent functional alterations in brain regions supporting cognitive control and reward processing | 2018 | Cannabis | Diagnostic | Hemodynamic | fMRI | Multiple |
| Neurobiological correlates of cue-reactivity in alcohol-use disorders: A voxel-wise meta-analysis of fMRI studies | 2021 | Cannabis | Diagnostic | Hemodynamic | fMRI | Cue-reactivity |
| Neurobiological correlates of cue-reactivity in alcohol-use disorders: A voxel-wise meta-analysis of fMRI studies | 2021 | Cannabis | Response | Hemodynamic | fMRI | Cue-reactivity |
| The differential neural substrates for reward choice under gain-loss contexts and risk in alcohol use disorder: Evidence from a voxel-based meta-analysis | 2023 | Alcohol | Diagnostic | Hemodynamic | fMRI | Loss and Gain |
| A meta-analysis of the neural substrates of monetary reward anticipation and outcome in alcohol use disorder | 2023 | Alcohol | Diagnostic | Hemodynamic | fMRI | MID |

| | | | | | | |
|---|---|---|---|---|---|---|
| A meta-analysis of the neural substrates of monetary reward anticipation and outcome in alcohol use disorder | 2023 | Alcohol | Diagnostic | Hemodynamic | fMRI | MID |
| The differential neural substrates for reward choice under gain-loss contexts and risk in alcohol use disorder: Evidence from a voxel-based meta-analysis | 2023 | Alcohol | Diagnostic | Hemodynamic | fMRI | Risky Decision-Making |
| Shared gray matter alterations in subtypes of addiction: a voxel-wise meta-analysis | 2021 | Cocaine/ Nicotine/ Alcohol | Diagnostic | Structural | sMRI | NA |
| Electrophysiological indexes for impaired response inhibition and salience attribution in substance (stimulants and depressants) use disorders: A meta-analysis | 2021 | General | Diagnostic | Electrophysiology | EEG | Cue-Reactivity |
| Electrophysiological indexes for impaired response inhibition and salience attribution in substance (stimulants and depressants) use disorders: A meta-analysis | 2021 | General | Response | Electrophysiology | EEG | Cue-Reactivity, Inhibition, Flanker |
| Voxelwise meta-analysis of gray matter anomalies in chronic cigarette smokers | 2016 | Nicotine | Diagnostic | Structural | sMRI | NA |

**References:**


1. Aggregate Analysis of ClinicalTrials.gov (AACT) Database. Clinical Trials Transformation Initiative (CTTI). https://aact.ctti-clinicaltrials.org

2. R Core Team. R: A language and environment for statistical computing. Published online 2013.

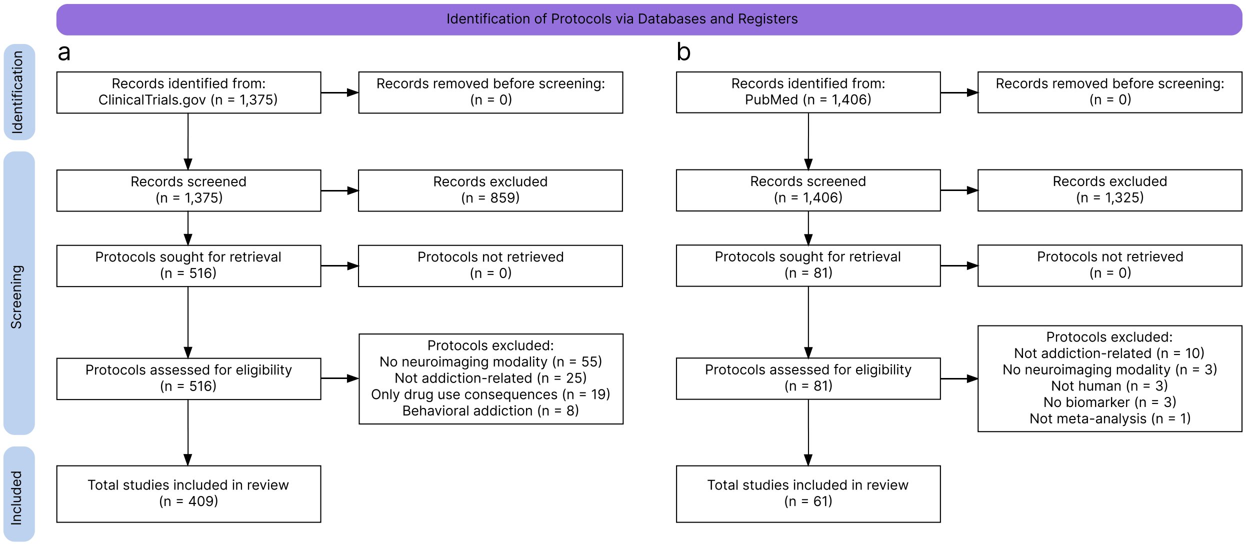


***Supplementary Figure 1: PRISMA2020 compliant flow diagrams.*** *A. Study protocols using putative neuroimaging biomarkers b. Meta-analyses providing evidence for putative neuroimaging biomarkers.*

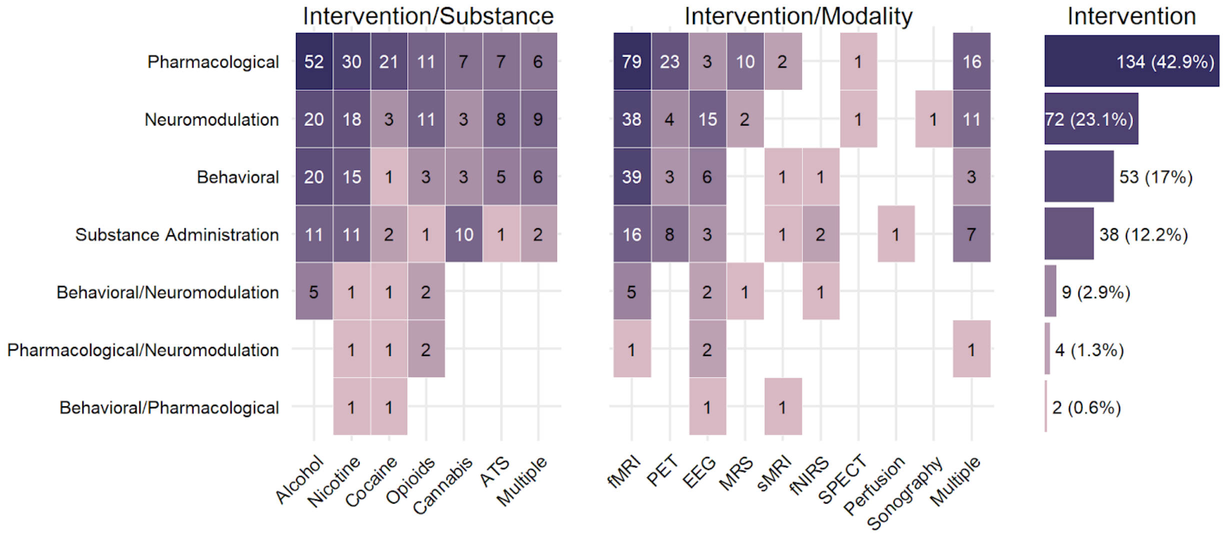


***Supplementary Figure 2:*** *Interventional protocols divided alternatively by the substance of interest and by neuroimaging modality used in the protocol database obtained from the systematic review in clinical trials.gov (n total = 312). The horizontally aligned bar plot presents the total number of protocols using each type of interventional manipulation.*

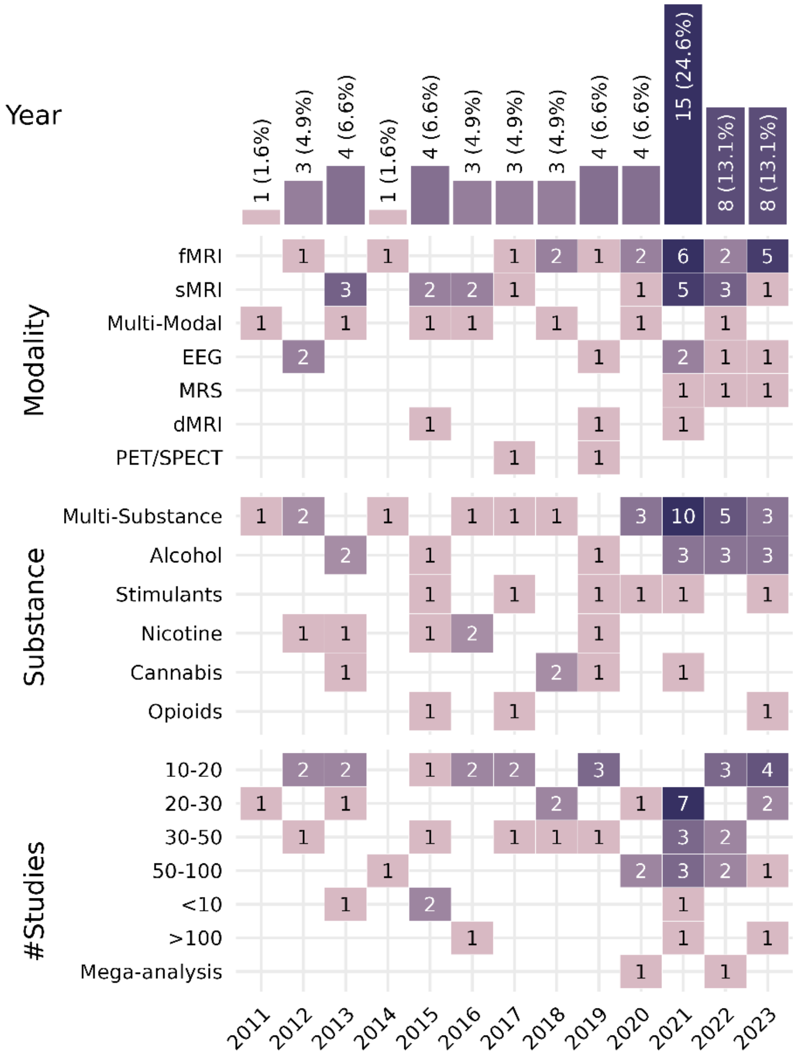


***Supplementary Figure 3: Breakdown of neuroimaging meta-analysis studies across years.*** *Number of included neuroimaging meta-analysis publications in each year, broken down by imaging modality, substance use disorder, and total number of studies included in the meta-analysis. sMRI: structural MRI, including whole-brain T1 imaging, gray matter volumetry, or diffusion tensor imaging. Perfusion: brain perfusion imaging, including arterial spin labeling, cerebral blood flow imaging, and magnetic resonance angiography. MRS: magnetic resonance-spectroscopy. Data were collected from ClinicalTrials.gov on November 17, 2021.*
